# Reclaiming Hand Functions for Chronic Spinal Cord Injury with Epidural Brain-Computer Interface

**DOI:** 10.1101/2024.09.05.24313041

**Authors:** Dingkun Liu, Yongzhi Shan, Liang Chen, Wenzheng Li, Xintong Yao, Ruwei Yao, Junze Zheng, Fangshuo Liang, Wen Zhu, Honglai Xu, Tao Liu, Yujing Wang, Penghu Wei, Yin Mao, Guoguang Zhao, Bo Hong

**Author notes:** Equal contribution (Dingkun Liu, Yongzhi Shan, Liang Chen).

## Abstract

Loss of hand function after cervical spinal cord injury limits independence in daily life. Implanted brain–computer interfaces (BCIs) can assist movement, but their role in sustained hand rehabilitation at home remains unclear^1,2^. Here we show that a wireless epidural BCI provides long-term hand assistance at home, accompanied by gains in unassisted hand function in two people with chronic cervical spinal cord injury. The system decoded grasp intention from electrodes placed above the dura to control a pneumatic glove, coupling motor intention to hand movement and sensory feedback. The system supported home use for 24 and 12 months, with 98% success in object-transfer testing and assistance during eating and drinking. Both participants regained the ability to grasp and hold objects without BCI assistance. Functional gains were observed in both hands despite unilateral training and were accompanied by increased somatosensory evoked responses. These findings suggest that sustained BCI-assisted hand use at home can support recovery of voluntary control after chronic spinal cord injury.

## Introduction

Cervical spinal cord injury can result in severe and lasting impairment of hand function, with major consequences for independence in daily life. Restoration of grasp and object manipulation remains a major unmet clinical need in persons with tetraplegia. Implanted neurotechnologies have shown increasing promise for the restoration of motor function after paralysis. Brain– computer interfaces can decode cortical activity to control external assistive devices, and restorative neuromodulation strategies can engage spared pathways and promote activity-dependent plasticity during rehabilitation^1–6^. Yet major translational challenges remain. Many current approaches depend on highly invasive neural interfaces, direct neuromodulatory interventions, or structured laboratory-based training paradigms, which may limit long-term safety, durability, and practicality for sustained use in daily life^7,8^.

More broadly, it remains unclear whether an implanted BCI system can be designed not only to assist impaired hand function, but also to support rehabilitation through repeated use over time. By translating grasp intention into movement of the impaired hand, BCI-controlled assistance can couple attempted movement with remained sensory feedback. Repeated pairing of motor intention with its sensory consequences may promote activity-dependent plasticity within spared sensorimotor networks, providing a potential basis for recovery of voluntary motor control^9,10^. Integrating this process into everyday grasping and object manipulation could create opportunities for repeated, task-specific practice beyond supervised rehabilitation sessions. Realizing this potential requires a system that provides reliable assistance and remains practical for sustained use at home. Longitudinal evaluation must therefore establish both its ability to enable hand function during use and whether repeated use is accompanied by functional gains that persist without device assistance.

A minimally invasive epidural interface may offer one such approach by reducing tissue disruption while preserving signals suitable for motor decoding. We developed a wireless, minimally invasive epidural BCI to decode grasp intention and control a wearable hand exoskeleton. In this study, we evaluated whether this system could enable assistive hand function and support recovery in patients with tetraplegia due to spinal cord injury. In two patients, the system supported home use for up to 24 months, with a 98% success rate in object-transfer testing and meaningful recovery of neurologic and hand function.

## Results

### Longitudinal Characteristics of Epidural Brain Signals

Usually, brain electrophysiological recordings can be divided into three distinct levels: scalp electroencephalography (EEG), subdural electrocorticography (sECoG), and intracortical single-unit recordings. These methods vary in invasiveness and signal quality, with EEG being the safest but having the lowest signal quality, and action potential recordings being the most invasive but offering the highest signal quality. We propose that epidural ECoG (eECoG) represents a fourth type, which provides higher safety than intracortical recordings^7^ and is considered minimally invasive. However, long-term eECoG recordings have been scarce. With NEO BCI system, we obtained substantial amount of eECoG signals for the first time. By analyzing these signals’ characteristics in spatial, temporal and spectral domains, we can maximize the use of eECoG signals in the design of NEO BCI decoding methods.

The time-frequency representations of eECoG recordings from two patients at 60–90 days post-surgery were analyzed (Fig. 1e). Specifically, P01 had 90 trials recorded, while P02 had 780 trials. The spectral patterns of both patients were similar to those observed with sECoG, exhibiting a characteristic event-related spectral perturbation (ERSP) pattern, with low-frequency power suppression (15–50 Hz) and high-frequency power enhancement (>50 Hz)^11,12^.

**Figure 1.**
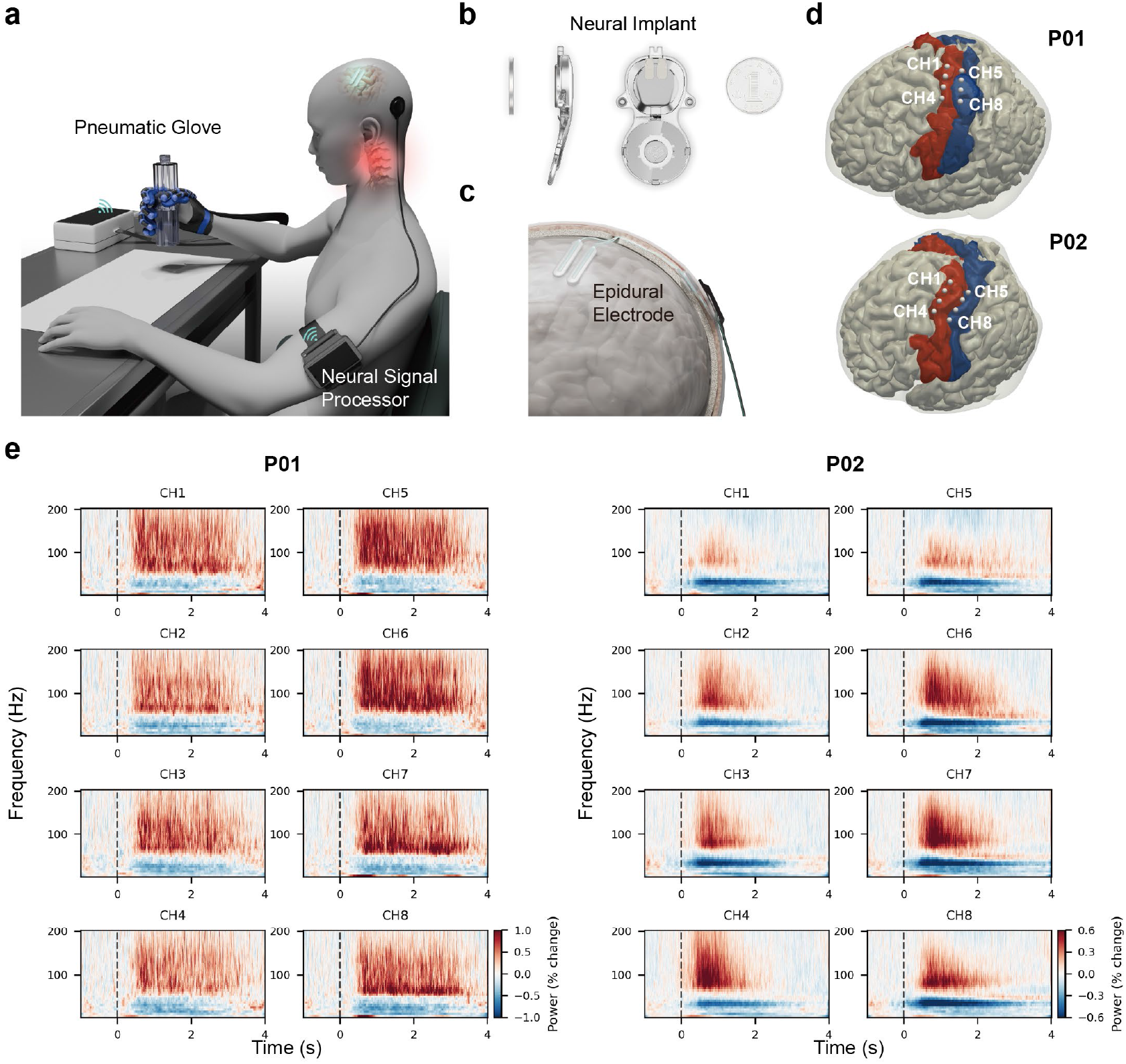
NEO system design and epidural signal characteristics. a, Diagram of the NEO brain-computer interface (BCI) system. The NEO BCI system transmits epidural ECoG signal through a coil to an external device attached to the patient’s arm. The external device sends the signals to the host computer, where an algorithm decodes the patient’s grasping intention and drives a wearable pneumatic glove to grasp objects. When the patient stops imagining the grasp, the pneumatic glove is driven to open and release the object. b, Structure of the NEO implant. The upper part is the implant, and the lower part is the internal coil. c, The NEO implant. Electrodes are fixed on the surface of the dura mater and connected to the implant through a skull tunnel. The external coil is magnetically attached through the skin, transmitting signals and power. d, Epidural electrode positions in two patients. CH1–CH4 are located on the precentral gyrus colored in red, while CH5–CH8 are positioned over the postcentral gyrus in blue. e, Percentage change of power during imagined grasping relative to rest for both patients. The spectral pattern exhibits a typical dual-frequency characteristic, including low-frequency (15-50 Hz) suppression and high- frequency (>50 Hz) enhancement.

We compared the spatio-temporal-spectral characteristics of P01’s epidural ECoG with subdural ECoG datasets^13^, the patient’s own scalp EEG, and a published EEG dataset^14^. Scalp EEG showed no significant event-related spectral perturbation (ERSP) above the low gamma (>40 Hz) frequency band (p=0.42, independent t-test, Fig. 2a). Both epidural and subdural ECoG showed a similar ERSP pattern, with low-frequency power suppression and high-frequency power enhancement. It is particularly noteworthy that the frequency upper limit of eECoG can exceed 200 Hz, which is significantly higher than that of scalp EEG (Fig. 2a). The power spectral density of eECoG was lower than that of sECoG in most frequency bands but obviously higher than that of scalp EEG in the high-frequency range (20-75 Hz) (Fig. 2b). In spatial domain, by comparing the relationship between channel correlation and electrode distance, we qualitatively verified the spatial resolution differences among these three types of recordings. The correlation between EEG channels decreased slowly with increasing electrode distance, indicating lowest spatial resolution. The correlation between sECoG channels decreased most rapidly with increasing electrode distance, indicating the highest spatial resolution, with eECoG showing moderate spatial resolution (Fig. 2c).

**Figure 2.**
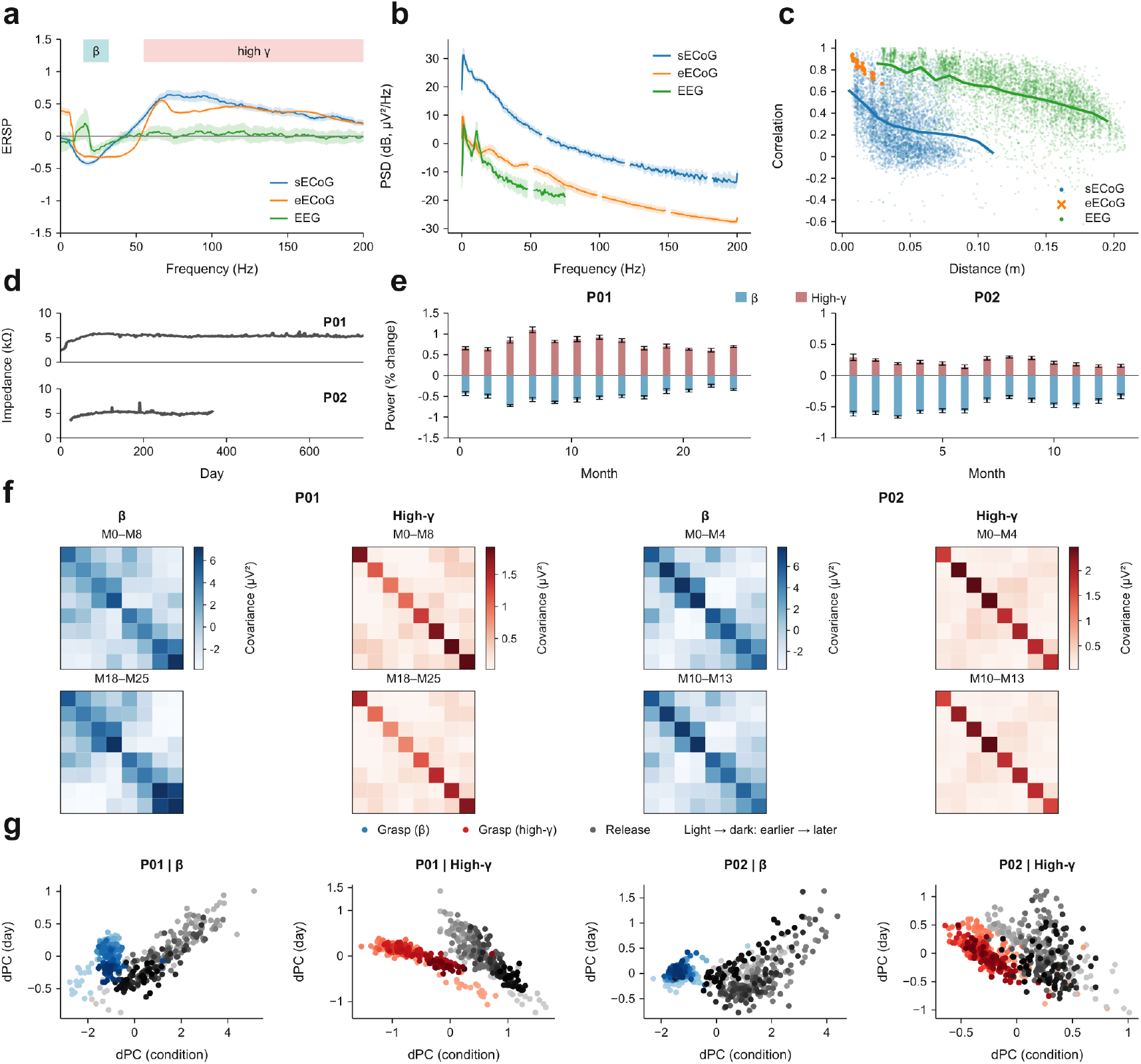
Long-term spatio-temporal-spectral characteristics of epidural signals. a, Comparison of event-related spectral perturbations (ERSP) among subdural ECoG, epidural ECoG, and scalp EEG. Epidural ECoG exhibits an effective frequency band range similar to subdural ECoG. b, Comparison of Power spectral density (PSD) among the three types of electrophysiological recordings. Subdural ECoG has the highest amplitude, and epidural ECoG has a higher amplitude in the higher frequency band (30-70 Hz) compared to scalp EEG. The PSD excludes the power line frequencies. c, Relationship between channel signal correlation and electrode distance. d, Impedance remained stable across the implantation period for both subjects. e, Grasp movement related power change of High-gamma and beta band across different months. f, Covariance matrices of High-gamma and beta band across different months, the spatial distribution of neural activity remains consistent. g, Demixed PCA (dPCA) of Riemannian manifold features. Multi-month neural features in the Riemannian tangent space were decomposed using dPCA to isolate task-related variance from temporal drift. The color gradients (from light to dark) represent the longitudinal progression across the study period (730 days for P01, 365 days for P02).

We continuously monitored epidural electrode impedance to assess postoperative intracranial environmental changes. Initial impedance showed a mild increase (P01 <600 Ω, P02 <300 Ω) and stabilized after 15 weeks (Fig. 2d). Concurrently, frequency-domain features of neural signals evolved over time: during grasping tasks, the extent of high-frequency enhancement and low-frequency suppression remained relatively stable over extended periods (730 days for P01, 365 days for P02) without significant fluctuation.

At the feature-space level, spatial covariance matrices of high- and low-frequency signals demonstrate consistent structural stability. Comparison of covariance matrices over up to 24 months (Fig. 2f) revealed that the underlying neural manifold associated with grasping intention maintained significant structural stability despite prolonged in-home use. To decouple the effect of temporal drift on task discrimination, we applied demixed principal component analysis (dPCA)^15^ to perform dimensionality reduction on features projected onto the Riemannian tangent space (Fig. 2g). The dPCA results showed that task-related structure (grasping vs. rest) remained clearly separated in state space, whereas temporal evolution (represented by the color gradient from light to dark) followed a different trajectory across the recording period. This finding suggests that despite subtle long-term neural signal drifts, the core task-discriminative features remain highly robust on the Riemannian manifold, supporting stable asynchronous BCI decoding without frequent recalibration.

### BCI-assisted Natural Grasping

The minimally invasive epidural BCI enabled reliable decoding of grasp intention and effective assistance of hand motor behavior. In a representative free-grasping session, single-trial epidural ECoG showed clear task-related spectral modulation with a high signal-to-noise ratio, and the decoder generated grasp probabilities that closely followed the participant’s intended grasping periods (Fig. 3a). These results demonstrate that grasp-related neural activity could be detected continuously and translated into real-time control signals for the wearable pneumatic glove.

**Figure 3.**
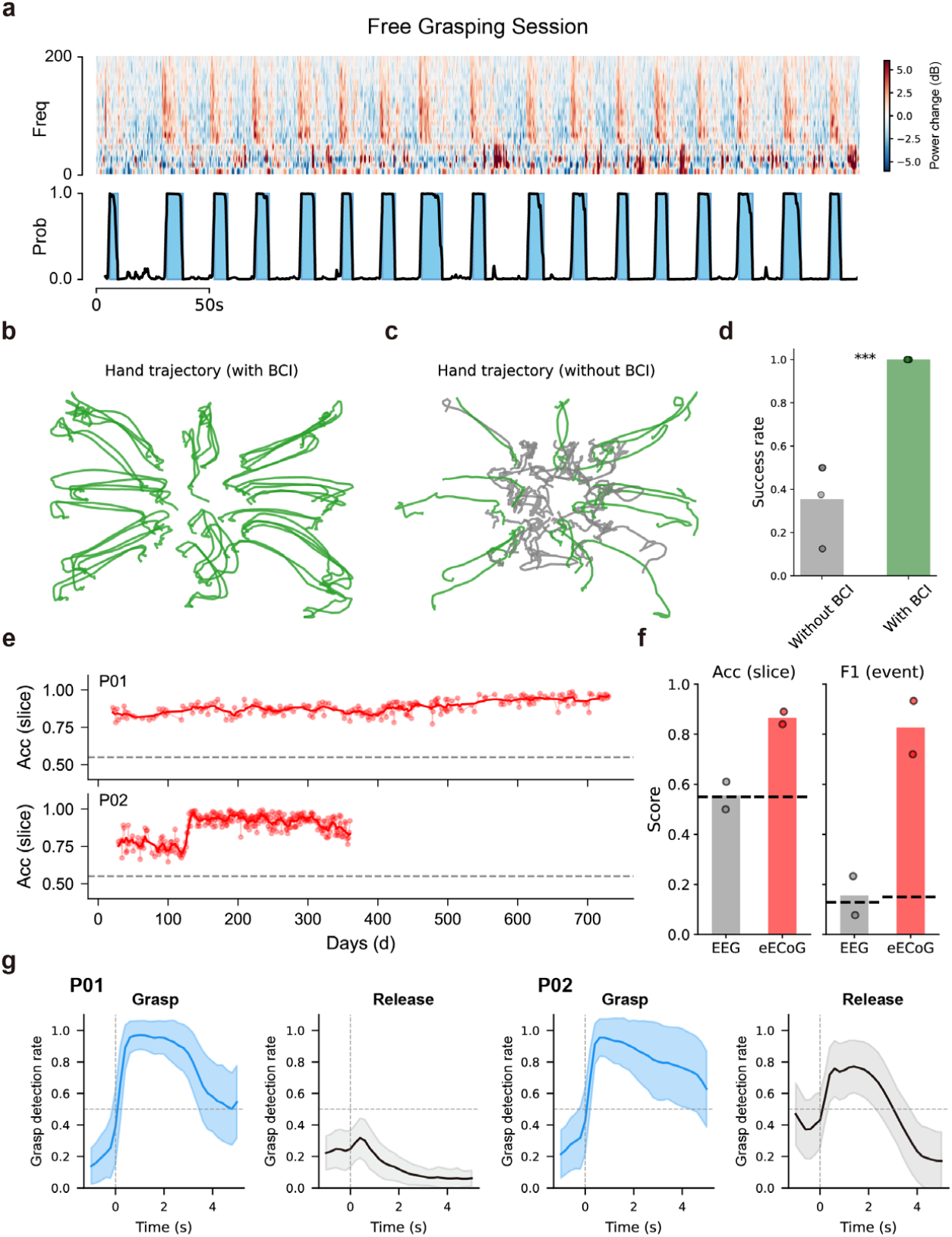
Minimally invasive epidural BCI for motor assistance. a, Example data from a free grasping session of P01. The upper panel shows the time–frequency spectrogram of single-trial epidural ECoG, demonstrating a high signal-to-noise ratio. The lower panel shows the grasping probability decoded in real time; blue shading marks the periods identified as grasping by the decoder. b,c, Hand movement trajectories of P01 during an object-moving task with BCI assistance (b) or unaided (c), recorded by a top-down camera and extracted using a keypoint detection model. Green lines denote successful trials; grey lines denote failed trials. d, Task success rates for P01 to move the object to the designated position within 10 seconds. With BCI assistance the patient achieved a 100% success rate, compared with 35% unaided (\*\*\**P* < 0.001). Each dot represents one session. e, Long-term stability of the offline slice-level decoding accuracy for two patients (P01, upper; P02, lower). Dots represent individual sessions; the solid line shows the 7-day running average. The grey dashed line indicates chance level (55%). Both patients maintained high accuracy over the full observation period (730 and 365 days, respectively). f, Comparison of slice-level accuracy (left) and event-level F1 score (right) between scalp EEG and epidural ECoG (eECoG). Each dot represents a different subject. Black dashed lines indicate chance levels. Epidural ECoG substantially outperformed scalp EEG on both metrics. g, Instantaneous grasp detection rate aligned to movement onset for P01 and P02. The left subplots show the rate at which the classifier outputs “grasp” during actual grasp events (blue curves); the right subplots show the same metric during release events (grey curves). Shaded regions indicate cross-trial variability (STD). The vertical dashed line marks movement onset; the horizontal dashed line marks 50%. The time-resolved detection profiles indicate clear separability of grasp and release states, with a slower return toward baseline after release in P02 than in P01.

We next evaluated whether BCI assistance improved functional performance in an object-transfer task. When assisted by the BCI system, participant P01 was able to grasp the object and move it from the center to the designated target location more effectively, with movement trajectories showing consistently successful transfers. In contrast, under unaided conditions, trajectories were more limited and failures were frequent (Fig. 3b, c). Quantitatively, BCI assistance markedly improved task success rate: P01 achieved a 100% (49/49) success rate for moving the object to the designated position within 10 s, compared with only 35% (17/48) without assistance (Fig. 3d, Table S4-S5). P02 also achieved 96% (46/48) success rate in object-transfer tasks (Table S6). These findings indicate that the epidural BCI provided substantial functional support for goal-directed hand actions.

To examine long-term robustness, we tracked offline slice-level decoding accuracy across sessions in both participants using a 500-msec sliding window, corresponding to the maximum delay for visual feedback in the continuous decoding framework (see Supplementary Appendix, Model Training and Evaluation). Decoding performance remained consistently above chance level throughout the full observation period, including up to 730 days in P01 and 365 days in P02 (Fig. 3e), demonstrating stable long-term performance of the minimally invasive epidural BCI system. Using the same decoding framework, epidural ECoG outperformed scalp EEG in both slice-level accuracy (86.5% vs. 55.5%; chance, 55%) and event-level F1 score (82.7% vs. 15.5%; chance, 14.9%) (Fig. 3f), supporting the greater utility of epidural signals for grasp detection. The mean grasp-detection delay relative to the screen cue was 1.30±0.21 seconds, reflecting a conservative event-confirmation threshold chosen to reduce erroneous detections during real-time control rather than the 500-msec maximum visual-feedback delay itself; even so, this delay remained shorter than the 2-to-5-second range typically reported for EEG-based motor-imagery BCIs^16–18^.

Finally, to examine the pattern of continuous decoding across the grasp–release process, classifier outputs were aligned to movement cue onset. In both participants, the instantaneous detection rate increased around grasp onset and remained higher during grasp than during release, indicating clear separability of the decoded states over time rather than a simple static binary classification. Individual differences were also evident, with a slower return toward baseline after release in P02 than in P01 (Fig. 3g).

In a home use setting, NEO BCI can assist with various tasks requiring grasping. However, activities like eating and drinking usually elicit significant electromyographic (EMG) noise, which interferes with the BCI decoder. Since the spatial distribution of EMG noise differs from that of grasping movements, the Riemannian geometry method, which is highly sensitive to spatial patterns, can filter out noise without supervised training (Fig. S11). Compared to the Riemannian geometry method, the Linear and CSP methods (see Appendix: Control Decoding Methods) had a false activation rate (FAR) of 100% during chewing, while the Riemannian geometry method had a FAR of only 20% (Fig. S11). Using the Riemannian geometry BCI decoder, the patient could perform grasp-related daily tasks such as eating and drinking (Video 5, 267 days after implantation), which was impossible before the BCI implantation.

### Hand Function Rehabilitation and Electrophysiology Assessment

We hypothesized that the coupling of top-down cortical activity with bottom-up peripheral sensory feedback during BCI-assisted grasping could promote neural reorganization at the spinal cord lesion site and thereby facilitate rehabilitation^19,20^. Consistent with this idea, both patients showed progressive neurophysiological and functional recovery during long-term training. Somatosensory evoked potentials (SEPs) elicited by stimulation of the radial, median and ulnar nerves provided a physiological readout of sensory pathway function at different spinal levels. Because the radial nerve mainly reflects segments above the zone of partial preservation, whereas the median nerve lies closer to it^21^, we expected the median nerve to show greater longitudinal change than the radial nerve.

This pattern was observed in both patients. After 24 months of training in P01, the peak amplitude of SEPs elicited by median nerve stimulation at CH7 increased by 13.1 μV, whereas the corresponding increase for the radial nerve was only 0.72 μV (Fig. 4b). A similar pattern was observed in P02 after 12 months of training, with the peak amplitude of SEPs elicited by median nerve stimulation at CH6 increasing by 23.34 μV, compared with only 4.44 μV for the radial nerve (Fig. 4b). Further analysis of the early (15-35 ms) and late (40-80 ms) SEP components (see Appendix: SEP Measurement) showed that, in both patients, responses elicited by MN stimulation underwent greater amplitude enhancement than those elicited by RN stimulation (Fig. 4c). Spatially, enhancement of both early and late components was concentrated mainly over electrodes corresponding to the postcentral gyrus, consistent with strengthened sensory cortical responses (Fig. 4c, Fig. S15-S16). Responses elicited by ulnar-nerve stimulation were detectable in both patients but were generally weaker and less prominent than those elicited by median-nerve stimulation. Modest longitudinal enhancement was observed at selected channels, with clearer changes in P01 and more limited, spatially sparse changes in P02 (Fig. S12-S16 and Table S3).

**Figure 4.**
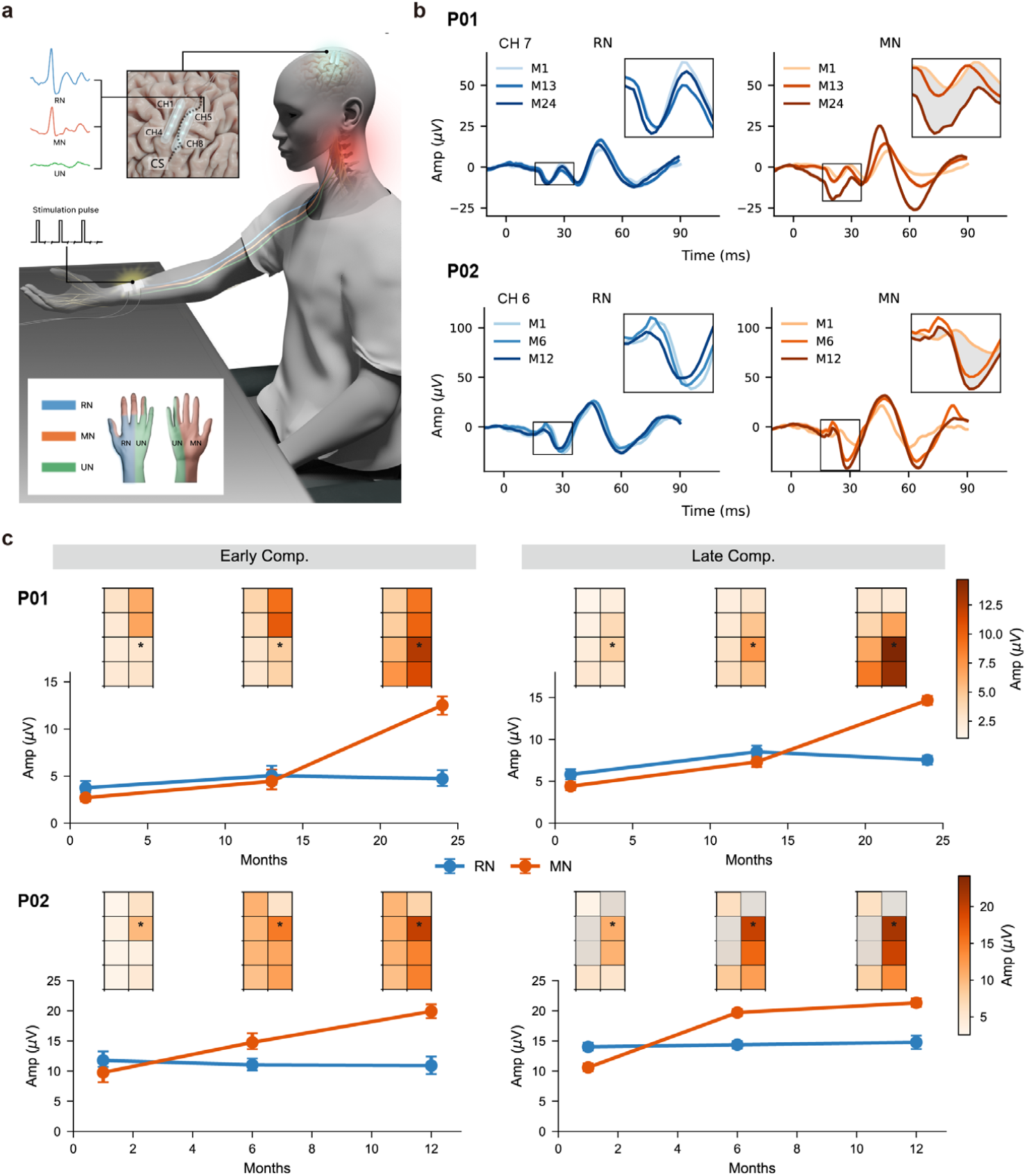
Electrophysiological rehabilitation assessments. a, Schematic of the sensory evoked potential (SEP) measurement setup. Transcutaneous stimulation of the patient’s radial (RN), median (MN), and ulnar (UN) nerves was delivered by functional stimulators, and SEP responses were recorded from the NEO epidural electrodes. b, Representative SEP waveforms of the RN (left) and MN (right) recorded from two patients at different months. For P01, recordings from Electrode No. 7 during the first, thirteenth, and twenty-fourth months are shown; for P02, recordings from Electrode No. 6 during the first, sixth, and twelfth months are shown. The insets show magnified views of specific latencies. The shaded areas indicate differences between the first month (M1) and the last month (M24 for P01 and M12 for P02). c, Changes in average amplitude over time for early (15-35 ms, left) and late (40-80 ms, right) SEP components of the RN and MN in two patients. The upper row shows P01, and the lower row shows P02. The 2 × 4 heatmaps above show the spatial distribution patterns of the corresponding components across eight electrodes; the line plots show changes in SEP component amplitude over months at the electrode indicated by the asterisk in the heatmaps. Results for UN are listed in the supplementary appendix (Fig. S12-S16).

The patients’ neurological scales and hand function assessments corresponded well with the SEP changes. ARAT scores increased progressively over the course of training in both patients (Fig. 5b-c, Table S7-S8). In patient P01, the ARAT score of the trained hand increased from 21 at baseline to 43 at the final assessment, while in P02, the score increased from 22 to 34 (Fig. 5c, red). Notably, although BCI training was applied only to one hand, the contralateral hand also showed substantial improvement of 13 and 18 points respectively (Fig. 5b-c, gray), demonstrating a bilateral functional gain. ISNCSCI assessment revealed concurrent improvements in both motor and sensory domains. In P01, the total motor score and sensory score increased by 16 and 12 points over 28 months, respectively (Fig. 5d). In P02, the total motor score and sensory score increased by 5 and 9 points over 6 months (Fig. 5e). In P01, a transient decline at month 7 coincided with baclofen and pregabalin use, after which scores recovered.

**Figure 5.**
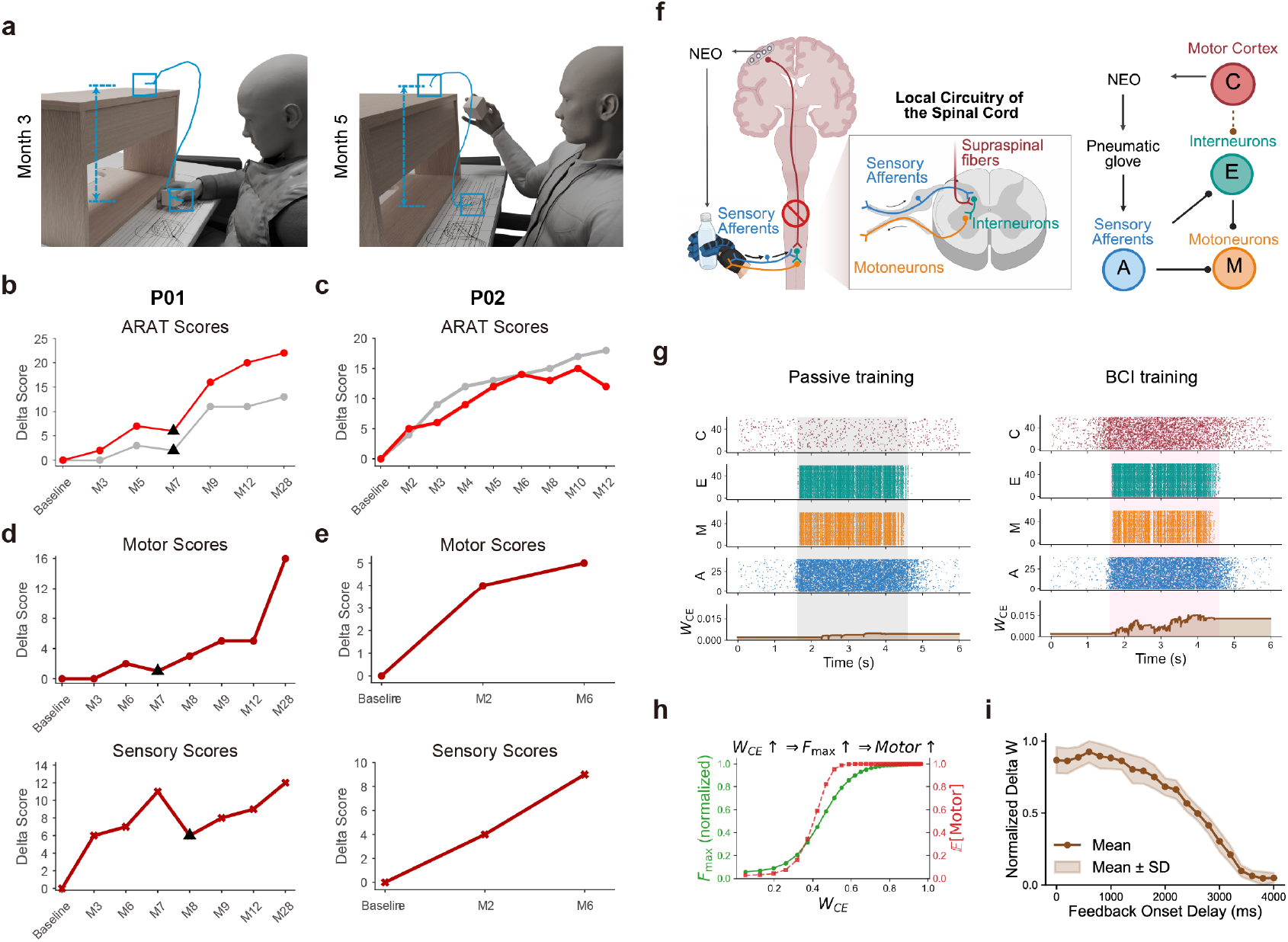
Assessments and dynamics of functional rehabilitation. a, A patient grasping and releasing a 5 cm block in the Action Research Arm Test (ARAT) at the 3rd and 5th months. The blue curve represents the block trajectory, the boxes mark the start and end points, and the dashed lines indicate the height of the lift. b-c, ARAT scores for patient P01 and P02, gray and red lines indicate left and right hand respectively. d-e, Changes in ISNCSCI total motor scores (upper) and sensory scores (lower) for P01 and P02, respectively. Motor and sensory scores encountered a decrease in the 7^th^ month (black triangles) due to the patient’s use of antispasmodic medication. f, Schematic overview of the C-E-M-A model representing the sensorimotor loop after spinal cord injury. Cortical activity (C) encodes grasp intention and projects to spinal interneurons (E), which in turn drive motoneurons (M). Afferent sensory feedback (A) is provided via a pneumatic glove. g, Raster plots of neural activity (C, E, M, A) and evolution of corticospinal synaptic strength (W_CE_) under two conditions. Left: passive training with glove-assisted grasp, where sensory input (A) drives spinal and motor activity in the absence of effective cortical drive. Right: BCI training, where imagined grasp (C) is temporally paired with glove-induced sensory feedback (A). h, Dependence of maximal force and expected motor test performance on corticospinal coupling strength (W_CE_). Normalized force (Fmax, green) and expected motor scores (red) are shown as nonlinear functions of W_CE_. i, Dependence of normalized corticospinal coupling strength (W_CE_) on feedback onset delay, showing the mean (solid line) and variability (shaded area, mean ± SD) across simulations.

### Modeling of Neural Circuits underlying BCI Training-Induced Functional Recovery

To investigate functional recovery, we modeled the corticospinal sensorimotor loop: primary motor cortex (M1) recruits spinal interneurons and α-motoneurons to generate muscle contraction^22,23^, while proprioceptive and cutaneous afferent feedback is integrated within spinal circuits^24–26^.

The reduced four-node model (C-E-M-A; Fig. 5f) comprises cortex (C), spinal interneurons (E), motoneurons (M), and afferent input (A)^27,28^. Corticospinal projections (C→E), initially weakened after injury, strengthen through spike-timing-dependent plasticity (STDP)^29–31^. Interneurons excite motoneurons (E→M), and afferent input modulates both populations (A→E and A→M). BCI-driven pneumatic glove actuation converts cortical activity into hand movement and sensory feedback, closing the loop.

During passive glove-assisted training, afferent input activated spinal interneurons and motoneurons, but corticospinal coupling strength (W_CE_) remained largely unchanged without coordinated cortical activity (Fig. 5g). BCI training temporally aligned cortical activity and sensory feedback, co-activating C and E and progressively increasing W_CE_. Stronger coupling elevated downstream motor activity and predicted increases in maximal force output (F_max_) and motor test performance (Fig. 5h)^32^.

Varying the delay between cortical activity and sensory feedback showed that synaptic strengthening (ΔW) was greatest at short delays (<800 ms), declined as the delay increased, and approached zero at longer latencies (Fig. 5i).

The model thus suggests that temporally precise coupling between motor intention and sensory feedback supports corticospinal strengthening, with low-latency feedback predicted to produce the greatest functional recovery.

## Discussion

In this first-in-human study, a wireless, minimally invasive epidural BCI provided durable assistive hand function and was associated with neurologic and functional improvement during prolonged home use. In two patients with tetraplegia due to cervical spinal cord injury, the system enabled reliable decoding of grasp intention, successful object transfer with a wearable exoskeleton, and performance of daily hand-related activities. During follow-up of up to 24 months, both patients recovered some ability to grasp and hold objects without BCI assistance, accompanied by improvements in ISNCSCI and ARAT scores and by increases in somatosensory evoked potentials, consistent with a corticospinal circuit model of activity-dependent functional recovery.

Implantable brain–computer interfaces must balance signal fidelity against surgical invasiveness and long-term durability. Microelectrode systems can provide high-resolution neural signals, but their chronic performance may be limited by tissue responses and the need for recalibration^8,33^. Subdural cortical interfaces have also shown sustained decoding performance, but they require closer contact with the brain surface and may carry greater procedural risk than epidural placement^7,33–35^. Our epidural system was designed for prolonged practical use through placement above the dura and a battery-free, wireless, closed-wound architecture. In two patients, this configuration supported home use for 36 months in total, with stable electrode impedance, preserved grasp-related neural signature, and sustained decoding performance across sessions. These findings suggest that epidural BCI recording may offer a clinically viable middle ground between highly invasive interfaces and long-term functional usability.

Beyond enabling assistive hand control, our findings suggest that repeated use of this BCI may have engaged a rehabilitative mechanism based on the repeated pairing of motor intention with sensory feedback. In both patients, improvement in unassisted grasping was accompanied by gains in ARAT and ISNCSCI scores and by increases in somatosensory evoked potentials, particularly for median-nerve stimulation, findings consistent with partial recovery of sensorimotor pathway function. Unlike previously reported brain-controlled rehabilitation systems that depend on direct electrical stimulation of the spinal cord or peripheral effectors^2,5,19,36–38^, the present approach coupled decoded cortical activity to glove-assisted hand movement, thereby generating reafferent sensory input without direct neurostimulation. The corticospinal circuit model provides a plausible mechanistic explanation for these observations: temporally coordinated activation of cortical and afferent inputs strengthened corticospinal coupling, whereas passive glove-assisted training alone produced little change. This increase in coupling predicts gains in force output and ARAT-like performance, and the effect was maximal at short feedback delays, supporting the view that low-latency closed-loop pairing between descending intent and sensory reafference is important for recovery after spinal cord injury.

This study has several limitations. It included only two participants and lacked a concurrent control condition, so the specific contribution of BCI-assisted training to the observed recovery cannot be determined. However, both participants were in the chronic stage of spinal cord injury at enrollment - more than 10 years after injury in P01 and 3 years after injury in P02 - whereas most spontaneous neurologic recovery after spinal cord injury occurs within the first 3 months and generally plateaus in the chronic stage^39^. In a clinical context in which effective options for restoring hand function remain limited, the functional and electrophysiological improvements observed here are therefore notable. Nonetheless, these findings should be interpreted cautiously and require confirmation in larger cohort studies. A larger clinical trial evaluating the safety and efficacy of the NEO system, conducted in accordance with Good Clinical Practice (GCP), has since been completed (NCT06990412).

Taken together, these results support the potential of a minimally invasive epidural BCI as a novel approach for restoring hand function and facilitating rehabilitation in chronic cervical spinal cord injury.

## Data Availability

All data produced in the present study are available upon reasonable request to the authors

https://zenodo.org/records/12818267

## Methods

### The Epidural Minimally Invasive BCI

The minimally invasive BCI, named NEO (Neural Electronics Opportunity), has an implant and an external processor that are linked with transcutaneous wireless communication. The exoskeleton hand (a pneumatic glove wore on the patient’s hand) is driven by the BCI external processor wirelessly as well (Fig. 1a). NEO includes two sets of coils: the first set supplies power for the implant through near-field induction, and the second transmits collected neural signals via Bluetooth to the external processor (Fig. 1c). As long as the external coil is magnetically attached on the skin and paired with the internal coil, the implant will be powered on. The implantable part of the NEO consists of a coin-size titanium alloy box, a coil, and epidural electrodes. The entire system is sealed with medical silicone material (Fig. 1b). Two epidural electrode strips are connected to the implant via electrode adaptors, with each electrode strip featuring four contacts capable of both recording and stimulating. These eight electrode contacts have a diameter of 3.2 mm and a center-to-center distance of 8 mm. The system operates at a sampling rate of 1 kHz.

This design has two advantages in reducing invasiveness. First, it employs minimally invasive epidural electrodes. These electrodes, placed on the dura mater, without penetrating the dura, prevent a strong immune response and ensure long-term stability. Second, the system utilizes bidirectional wireless communication for both power and signal transmission (Fig. 1c). Wireless communication reduces the risk of infection by eliminating the need for open wounds. The implant does not contain a battery; instead, it is powered wirelessly via the coils, which also handle bidirectional data transmission. This battery-free design ensures that the device’s lifespan is not limited by battery life. While a similar design has been widely adopted in cochlear implants^40^, similar concepts remain not widely adopted in implanted BCIs^41,42^.

Signals are received by a relay station attached to the arm and transmitted to a computer wirelessly. The computer decodes the patient’s motor control intention continuously from the neural data and drives the external pneumatic glove (Fig. 1a, Video 1). When the patient imagines grasping and holding, the pneumatic glove grasps; when the patient wants to release the object, the glove extends to release. In this unique configuration, the pneumatic glove not only acts as a BCI actuator but also provides sensory feedback to those neural fibers in the hand that may relay them to the injured spinal cord sites. Thus, in every trial of BCI hand control, the intrinsic top-down neural activities have a chance to meet with this bottom-up sensory feedback generated by the successful grasping and releasing.

### Study Participant

This study is part of a clinical trial for the implantable closed-loop BCI system (NEO) (ClinicalTrials.gov, NCT05920174), which was approved by the Xuanwu Hospital Ethics Committee in Apr. 2023 and by the Huashan Hospital Ethics Committee in Sept. 2024. Patient 01 (P01) was recruited in his 50s and had suffered a spinal cord injury in a car accident more than 10 years earlier, resulting in severely impaired bimanual motor function and making grasping, holding, and pinching actions impossible (Video 2). However, limited right arm function was preserved, allowing him to raise his right forearm within a constrained range of motion. Prior to NEO implantation, neurological assessments showed complete C4 spinal cord injury (AIS-A, Table S1, Fig. S1). Patient 02 (P02) was recruited in his 40s and had sustained a spinal cord injury 3 years earlier, with a similar pattern of motor impairment, although his residual motor function was slightly better. Neurological assessment before implantation showed incomplete C7 spinal cord injury (AIS-B, Table S2, Fig. S1). Informed consent was obtained from both participants.

### Implantation of Minimally Invasive BCI

To obtain the most informative neural signals related to hand grasping with a minimal number of electrodes, we performed preoperative planning for the epidural electrode implantation using functional MRI (fMRI). Research indicates that the transmission of ascending signals is crucial for SCI rehabilitation^43^. Therefore, our paradigm included fMRI imaging of both active and passive grasping attempts. The active grasping paradigm requires the subject to perform motor imagery based on visual cues, whereas the passive grasping paradigm requires the subject to perceive the sensation of the experimenter assisting him in grasping and releasing. The active grasping paradigm localized the sensory-motor area responsible for the patient’s hand, while the passive grasping paradigm assessed the residual ascending sensory signals (Fig. S2). Additionally, we collected CT and structural MRI images of the patient to determine the precise implantation site.

The significant activation regions were projected from the fMRI onto a 3D model of the cortex, using 3D modeling software to simulate electrode placement (Fig. S2). The position of the implant was determined using skull structure obtained from CT images, selecting an area with suitable thickness that can bury the titanium processor. The final plan was then output to a surgical navigation system to guide the implantation procedure.

The minimally invasive BCI implantation was conducted under general anesthesia. A coronal incision was made on the left scalp to place the internal processor, coil, and electrode strips. Following the preoperative plan, a circular window was created in the skull, exposing the dura mater, and the electrodes were sutured onto the dura mater according to the planned coordinates. The bone flap was then repositioned and secured. A 3-4 mm deep groove was created on the skull surface to embed the titanium implant, which was fixed using bone screws, and the incision was closed. The patient was discharged within 24 hours after all clinical observations were completed. No serious device-related or procedure-related adverse events occurred during implantation or follow-up (Table S9).

Postoperatively, we extracted the electrode positions from the postoperative CT images and registered them to the preoperative MRI images, obtaining a map of the eight electrode positions. For both patients, the first four electrodes were positioned over the precentral gyrus, while the latter four were placed over the postcentral gyrus. The anatomical parcellation of the electrodes was labeled based on the HCP-mmp1 atlas^44^. Specifically, channels CH1–CH4 correspond to HCP-MMP1.0 / area 6d, and channels CH5–CH8 correspond to HCP-MMP1.0 / area 1; these regions encompassed the most prominently activated hand areas identified by fMRI during hand movement imagination (Fig. 1d, Fig. S2).

### Decoding Brain-controlled Grasping Events

Based on the analysis of recorded epidural electrocorticography (eECoG) data (see Results: Long- term Characteristics of Epidural Signals), we confirmed that eECoG contains informative spatio- spectral features for decoding grasp intention. To accurately decode natural grasping movement intention, our BCI decoder integrated frequency, space, and time domain information, named GEMS (Geometry-Enhanced Multi-Spectral Model).

First, eECoG features were encoded as multi-band spatial covariance matrices, allowing the decoder to integrate spectral information across frequency bands with the intrinsic geometry of covariance representations for Riemannian classification^45,46^. Frequency bands were optimized for each participant based on calibration data and were typically selected from the beta, low-gamma, and high-gamma ranges; the final band selections for P01 and P02 are provided in the Supplementary Appendix (Fig. S5). Second, a hidden Markov model (HMM) was used to capture temporal dependencies between grasping and resting states, thereby improving the stability of continuous decoding (Fig. S6). The classifier was initialized using calibration-session data, and HMM transition parameters were estimated from uncued free-grasp recordings; the decoder was updated during longitudinal follow-up using calibration-session data as needed.

During longitudinal follow-up, decoder performance was assessed in cue-locked daily evaluation sessions (see Appendix: Brain–Computer Interface Training and Evaluation Paradigms).

Performance was evaluated at two complementary levels: accuracy for classifying short time windows of neural activity (slice-level), and F1 score for detecting grasp events within a predefined interval around the cued movement onset (event-level) (see Appendix: Model Training and Evaluation). These measures were intended to reflect, respectively, instantaneous decoding performance and practical continuous-control performance.

### Assessment of functional and neurological recovery

To assess the potential rehabilitative effects of the intervention, we performed electrophysiological measurements and longitudinal clinical evaluations. During training, voluntary movement intention generated descending cortical activity, whereas glove-assisted grasping generated ascending sensory input through intact peripheral afferent pathways; these two streams were expected to converge at the spinal cord lesion site. The intervention therefore paired cortical intention with peripheral reafference in a closed loop, unlike approaches based primarily on spinal stimulation^2,36,47^ and assistive BCI systems intended mainly for direct control of external devices without engaging a sensorimotor feedback loop^6,48^.

Somatosensory evoked potentials (SEPs) are used to assess sensory information conduction. SEPs are elicited by electrically stimulating peripheral nerves, and the resulting neural activity travels through the sensory pathways to the corresponding areas of the brain, such as the precentral and postcentral gyri. Thus, SEPs reflect the transmission of sensory information from peripheral nerves through the spinal cord to the brain, making them a useful tool for evaluating the extent of spinal cord injury and the progress of recovery^49,50^. SEP testing involves stimulating the median, radial, and ulnar nerves of the patient’s right arm (corresponding to C6-T1, C5-C8, and C8-T1, respectively) with a constant current and recording the SEPs from the precentral and postcentral gyri using the NEO epidural electrodes (Fig. 4a).

In this study, we employed two key assessments to quantify the neurological and functional recovery of the subjects. The international standards for neurological classification of spinal cord injury (ISNCSCI) scale^51^ was used to evaluate the overall neurological status, including motor and sensory capabilities, which is vital for determining the extent of spinal cord damage and subsequent recovery. Additionally, the action research arm test (ARAT) scale^52^ was utilized to assess the finer aspects of hand function, focusing on the subject’s ability to handle objects, which is crucial for evaluating the recovery of subtle motor skills of the upper limbs. These scales provided a quantifiable measure for monitoring the improvements in neurological function and hand dexterity over the course of the study.

## Acknowledgements

We thank our volunteer patients and their family for their commitment and trust. We thank Sichang Chen and Lin Liu from Xuanwu Hospital of Capital Medical University, Zehan Wu from Huashan Hospital, Fudan University, Xiaoshan Huang, Minchao Qi, Zheng Yan from Neuracle Technology Co., Ze’ao Xiong, Yunjing Li, Yujing Lin, Le Yu, Yichao Li from Tsinghua University, and Zhouxingyu Yan from Johns Hopkins University for their help in this work.

## Video 1 Mind to motion: NEO BCI overview

This video outlines the basic setup of the NEO brain-computer interface (BCI) system. First, the NEO implant is powered on. Next, the pneumatic glove is attached and configured. Once connected to the controller, the NEO system transmits epidural ECoG signals in real-time. Finally, subjects are able to control the pneumatic hand using his thoughts. Link to view online: https://cloud.tsinghua.edu.cn/f/ba945e673ec74af5a2b5/

## Video 2 P01 ARAT hand function Test

This video demonstrates P01’s improvement in hand function following a 9-month training period. It compares the subject’s performance on ARAT tasks before surgery (baseline) and at the 9-month mark for both hands. For the right hand, grasping tasks involving 5 cm, and 7.5 cm cubes, as well as a 7.5 cm sphere, are compared between the baseline and 9^th^ month. For the left hand, the grasping task with a sharpening stone is evaluated. Link to view online: https://cloud.tsinghua.edu.cn/f/d6eaeb7ac139458c9055/

## Video 3 P02 ARAT hand function Test

This video demonstrates the longitudinal change in ARAT performance in patient P02. It compares task performance before surgery (baseline) and after training for both hands. For the right hand, grasping tasks involving a 10 cm wooden block, a 7.5 cm-diameter sphere, and a 1 cm-diameter cube are compared between baseline and the 4th, 5th, and 10th postoperative months, respectively. For the left hand, grasping tasks involving a 10 cm wooden block and a 7.5 cm-diameter sphere are compared between baseline and the 4th and 10th postoperative months, respectively. Link to view online: https://cloud.tsinghua.edu.cn/f/e6cc70b6606146e7ab5c/

## Video 4 P01 NEO BCI for grasping and object moving tasks

This video demonstrates how the NEO system assists with hand function. With the help of the NEO BCI system, the subject successfully lifts a bottle of water and moves it to the designated target area. Link to view online: https://cloud.tsinghua.edu.cn/f/ec097b01744348dca7e1/

## Video 5 NEO for Independent Home Use

This video highlights how the NEO system enhances the subject’s daily life. Using the NEO system, the subject can independently eat with a customized fork and drink from either a plastic bottle or glass. Link to view online: https://cloud.tsinghua.edu.cn/f/380a1549ac39487d8895/

## References

1. Benabid, A. L. et al. An exoskeleton controlled by an epidural wireless brain–machine interface in a tetraplegic patient: a proof-of-concept demonstration. Lancet Neurol. 18, 1112–1122 (2019).

2. Lorach, H. et al. Walking naturally after spinal cord injury using a brain–spine interface. Nature 618, 126–133 (2023).

3. Moses, D. A. et al. Neuroprosthesis for Decoding Speech in a Paralyzed Person with Anarthria. N. Engl. J. Med. 385, 217–227 (2021).

4. Samejima, S. et al. Brain-computer-spinal interface restores upper limb function after spinal cord injury. IEEE Trans. Neural Syst. Rehabil. Eng. 29, 1233–1242 (2021).

5. Kilgard, M. P. et al. Closed-loop vagus nerve stimulation aids recovery from spinal cord injury. Nature 643, 1030–1036 (2025).

6. Willett, F. R., Avansino, D. T., Hochberg, L. R., Henderson, J. M. & Shenoy, K. V. High-performance brain-to-text communication via handwriting. Nature 593, 249–254 (2021).

7. Branco, M. P., Geukes, S. H., Aarnoutse, E. J., Ramsey, N. F. & Vansteensel, M. J. Nine decades of electrocorticography: A comparison between epidural and subdural recordings. Eur. J. Neurosci. 57, 1260–1288 (2023).

8. Patel, P. R. et al. Utah array characterization and histological analysis of a multi-year implant in non-human primate motor and sensory cortices. J. Neural Eng. 20, 014001 (2023).

9. Mrachacz-Kersting, N. et al. Efficient neuroplasticity induction in chronic stroke patients by an associative brain-computer interface. J. Neurophysiol. 115, 1410–1421 (2016).

10. Biasiucci, A. et al. Brain-actuated functional electrical stimulation elicits lasting arm motor recovery after stroke. Nat. Commun. 9, 2421 (2018).

11. Miller, K. J. et al. Spectral changes in cortical surface potentials during motor movement. J. Neurosci. 27, 2424–2432 (2007).

12. Bundy, D. T. et al. Characterization of the effects of the human dura on macro- and micro-electrocorticographic recordings. J. Neural Eng. 11, 016006 (2014).

13. Miller, K. J. A library of human electrocorticographic data and analyses. Nat. Hum. Behav. 3, 1225–1235 (2019).

14. Schalk, G., McFarland, D. J., Hinterberger, T., Birbaumer, N. & Wolpaw, J. R. BCI2000: A General-Purpose Brain-Computer Interface (BCI) System. IEEE Trans. Biomed. Eng. 51, 1034–1043 (2004).

15. Kobak, D. et al. Demixed principal component analysis of neural population data. eLife 5, e10989 (2016).

16. Pfurtscheller, G., Linortner, P., Winkler, R., Korisek, G. & Müller-Putz, G. Discrimination of motor imagery-induced EEG patterns in patients with complete spinal cord injury. Comput. Intell. Neurosci. 2009, 104180 (2009).

17. Ince, N. F., Arica, S. & Tewfik, A. Classification of single trial motor imagery EEG recordings with subject adapted non-dyadic arbitrary time-frequency tilings. J. Neural Eng. 3, 235–244 (2006).

18. Lawhern, V. J. et al. EEGNet: A compact convolutional neural network for EEG-based brain-computer interfaces. J. Neural Eng. 15, 056013 (2018).

19. Anderson, M. A. et al. Natural and targeted circuit reorganization after spinal cord injury. Nat. Neurosci. 25, 1584–1596 (2022).

20. Asboth, L. et al. Cortico–reticulo–spinal circuit reorganization enables functional recovery after severe spinal cord contusion. Nat. Neurosci. 21, 576–588 (2018).

21. Grisolia, J. S. & Wiederholt, W. C. Short latency somatosensory evoked potentials from radial, median and ulnar nerve stimulation in man. Electroencephalogr. Clin. Neurophysiol. 50, 375– 381 (1980).

22. Lemon, R. N. Descending Pathways in Motor Control. Annu. Rev. Neurosci. 31, 195–218 (2008).

23. Porter, R. & Lemon, R. Corticospinal Function and Voluntary Movement. (Oxford University Press, Oxford, 1995). doi:10.1093/acprof:oso/9780198523758.001.0001.

24. Pearson, K. G. Neural Adaptation in the Generation of Rhythmic Behavior. Annu. Rev. Physiol. 62, 723–753 (2000).

25. Proske, U. & Gandevia, S. C. The Proprioceptive Senses: Their Roles in Signaling Body Shape, Body Position and Movement, and Muscle Force. Physiol. Rev. 92, 1651–1697 (2012).

26. Azim, E. & Seki, K. Gain control in the sensorimotor system. Curr. Opin. Physiol. 8, 177–187 (2019).

27. Capogrosso, M. et al. A Computational Model for Epidural Electrical Stimulation of Spinal Sensorimotor Circuits. J. Neurosci. 33, 19326–19340 (2013).

28. Balaguer, J.-M. et al. Neural mechanisms underlying the recovery of voluntary control of motoneurons after paralysis with spinal cord stimulation. Neuron 113, 3846-3862.e6 (2025).

29. Nishimura, Y., Perlmutter, S. I., Eaton, R. W. & Fetz, E. E. Spike-Timing-Dependent Plasticity in Primate Corticospinal Connections Induced during Free Behavior. Neuron 80, 1301–1309 (2013).

30. Feldman, D. E. The Spike-Timing Dependence of Plasticity. Neuron 75, 556–571 (2012).

31. Dan, Y. & Poo, M. Spike Timing-Dependent Plasticity of Neural Circuits. Neuron 44, 23–30 (2004).

32. Fuglevand, A. J., Winter, D. A. & Patla, A. E. Models of recruitment and rate coding organization in motor-unit pools. J. Neurophysiol. 70, 2470–2488 (1993).

33. Milekovic, T. et al. Stable long-term BCI-enabled communication in ALS and locked-in syndrome using LFP signals. J. Neurophysiol. 120, 343–360 (2018).

34. Luo, S. et al. Stable Decoding from a Speech BCI Enables Control for an Individual with ALS without Recalibration for 3 Months. Adv. Sci. 10, 2304853 (2023).

35. Fountas, K. N. & Smith, J. R. Subdural electrode-associated complications: A 20-year experience. Stereotact. Funct. Neurosurg. 85, 264–272 (2007).

36. Moritz, C. et al. Non-invasive spinal cord electrical stimulation for arm and hand function in chronic tetraplegia: a safety and efficacy trial. Nat. Med. 30, 1276–1283 (2024).

37. Donati, A. R. C. et al. Long-Term Training with a Brain-Machine Interface-Based Gait Protocol Induces Partial Neurological Recovery in Paraplegic Patients. Sci. Rep. 6, 30383 (2016).

38. Jovanovic, L. I. et al. Restoration of Upper Limb Function after Chronic Severe Hemiplegia: A Case Report on the Feasibility of a Brain-Computer Interface-Triggered Functional Electrical Stimulation Therapy. Am. J. Phys. Med. Rehabil. 99, e35–e40 (2020).

39. Fawcett, J. W. et al. Guidelines for the conduct of clinical trials for spinal cord injury as developed by the ICCP panel: spontaneous recovery after spinal cord injury and statistical power needed for therapeutic clinical trials. Spinal Cord 45, 190–205 (2007).

40. Zeng, F.-G., Rebscher, S., Harrison, W., Sun, X. & Feng, H. Cochlear Implants: System Design, Integration, and Evaluation. IEEE Rev. Biomed. Eng. 1, 115–142 (2008).

41. Schalk, G. et al. Toward a fully implantable ecosystem for adaptive neuromodulation in humans: Preliminary experience with the CorTec BrainInterchange device in a canine model. Front. Neurosci. 16, 932782 (2022).

42. Ayyoubi, A. H. et al. Benchmarking signal quality and spatiotemporal distribution of interictal spikes in prolonged human iEEG recordings using CorTec wireless brain interchange. Sci. Rep. 14, 2652 (2024).

43. Formento, E. et al. Electrical spinal cord stimulation must preserve proprioception to enable locomotion in humans with spinal cord injury. Nat. Neurosci. 21, 1728–1741 (2018).

44. Glasser, M. F. et al. A multi-modal parcellation of human cerebral cortex. Nature 536, 171–178 (2016).

45. Congedo, M., Barachant, A. & Bhatia, R. Riemannian geometry for EEG-based brain-computer interfaces; a primer and a review. Brain Comput. Interfaces 4, 155–174 (2017).

46. Yger, F., Berar, M. & Lotte, F. Riemannian Approaches in Brain-Computer Interfaces: A Review. IEEE Trans. Neural Syst. Rehabil. Eng. 25, 1753–1762 (2017).

47. Inanici, F., Brighton, L. N., Samejima, S., Hofstetter, C. P. & Moritz, C. T. Transcutaneous Spinal Cord Stimulation Restores Hand and Arm Function after Spinal Cord Injury. IEEE Trans. Neural Syst. Rehabil. Eng. 29, 310–319 (2021).

46. Pandarinath, C. et al. High performance communication by people with paralysis using an intracortical brain-computer interface. eLife 6, e18554 (2017).

47. Kramer, J. K., Taylor, P., Steeves, J. D. & Curt, A. Dermatomal somatosensory evoked potentials and electrical perception thresholds during recovery from cervical spinal cord injury. Neurorehabil. Neural Repair 24, 309–317 (2010).

58. Kramer, J. L. K., Moss, A. J., Taylor, P. & Curt, A. Assessment of posterior spinal cord function with electrical perception threshold in spinal cord injury. J. Neurotrauma 25, 1019–1026 (2008).

49. Rupp, R. et al. International standards for neurological classification of spinal cord injury. Top. Spinal Cord Inj. Rehabil. 27, 1–22 (2021).

50. Yozbatiran, N., Der-Yeghiaian, L. & Cramer, S. C. A standardized approach to performing the action research arm test. Neurorehabil. Neural Repair 22, 78–90 (2008).

