## Supplementary Information for "Reclaiming Hand Functions for Chronic Spinal Cord Injury with Epidural Brain-Computer Interface"

FIGURE S2 FMRI LOCALIZATION TASK, FUNCTIONAL MAPPING, AND SURGICAL PLANNING IN

TWO PATIENTS (P01 AND P02). 22
FIGURE S3 LONGITUDINAL BAND-POWER CHANGES IN P01 AND P02. 23
FIGURE S4 SPATIAL DISTRIBUTION OF BASELINE HIGH-GAMMA ACTIVITY ACROSS MONTHS IN THE TWO PATIENTS. 24
FIGURE S5 FREQUENCY-BAND CLASS DISTINCTIVENESS IN THE TWO PATIENTS. 25
FIGURE S6 GEMS (GEOMETRY-ENHANCED MULTI-SPECTRAL) DECODING PIPELINE. 26
FIGURE S7 LONGITUDINAL DECODING PERFORMANCE IN PATIENT 1 ACROSS SEVEN MONTHS. 27
FIGURE S8 COMPARISON OF CONTINUOUS DECODING WITH AND WITHOUT HMM MODELING. 28
FIGURE S9 CALIBRATION AND TRAINING PARADIGMS. 29
FIGURE S10 LONGITUDINAL EVOLUTION OF EPIDURAL ECOG FEATURE DISTRIBUTIONS. 30
FIGURE S11 CHARACTERIZATION OF EMG NOISE IN DIFFERENT FEATURE SPACES. 31
FIGURE S12 LONGITUDINAL SOMATOSENSORY EVOKED POTENTIALS IN TWO PATIENTS. 32
FIGURE S13 LONGITUDINAL TRIAL-BY-TRIAL MAPS OF SOMATOSENSORY EVOKED POTENTIALS FOR PATIENT 01 (P01). 33
FIGURE S14 LONGITUDINAL TRIAL-BY-TRIAL SOMATOSENSORY EVOKED RESPONSES IN P02. 34
FIGURE S15 CHANGES IN AVERAGE SEP COMPONENT AMPLITUDE OVER TIME OF P01. 35
FIGURE S16 CHANGES IN AVERAGE SEP COMPONENT AMPLITUDE OVER TIME OF P02. 36
FIGURE S17 PNEUMATIC HAND KEYPOINT DETECTION MODEL. 37
FIGURE S18 TRAINING DURATION IN THE TWO PARTICIPANTS FOR A COMBINED 36 MONTHS BCI TRAINING. 38

**SUPPLEMENTAL TABLES** 39

TABLE S1 ISNCSCI SCORE OF P01 39
TABLE S2 ISNCSCI SCORE OF P02 40
TABLE S3 P-VALUES FOR SEP COMPONENT CHANGES IN P01 AND P02 41
TABLE S4 BCI-ASSISTED OBJECT TRANSFER SUCCESS RATE FOR P01 42
TABLE S5 UNAIDED OBJECT TRANSFER SUCCESS RATE FOR P01 42
TABLE S6 BCI-ASSISTED OBJECT TRANSFER SUCCESS RATE FOR P02 42
TABLE S7 ARAT SCORES OF P01 43
TABLE S8 ARAT SCORES OF P02 44
TABLE S9 ADVERSE EVENTS REPORTED DURING THE 1-YEAR FOLLOW-UP PERIOD 45

**REFERENCE** 46

### List of investigators and contributions

**List of investigators (authors):**

1. Dingkun Liu*
2. Yongzhi Shan*
3. Liang Chen*
4. Wenzheng Li
5. Xintong Yao
6. Ruwei Yao
7. Junze Zheng
8. Fangshuo Liang
9. Wen Zhu
10. Honglai Xu
11. Tao Liu
12. Yujing Wang
13. Penghu Wei
14. Yin Mao
15. Guoguang Zhao
16. Bo Hong

* These authors contributed equally

### Supplemental Text

#### S1 – Experimental procedures

##### S1.01 – Clinical Protocol and Study Participant

This study was part of a clinical trial of the implantable closed-loop brain-computer interface system (NEO) (ClinicalTrials.gov, NCT05920174). The trial was approved by the Ethics Committee of Xuanwu Hospital of Capital Medical University in April 2023 and by the Ethics Committee of Huashan Hospital in September 2024. Surgeries were performed at Xuanwu Hospital, while related assessments and rehabilitation were conducted at Tsinghua University and at the participants' homes. The overall study protocol included preoperative functional screening, surgical implantation, a calibration and adjustment period, and subsequent BCI-assisted active rehabilitation training. The rehabilitation schedule was individualized according to each participant's progress and health condition, and the corresponding daily usage is summarized in Fig. S18.

Two participants with cervical spinal cord injury were enrolled in this study. Patient 01 (P01) was a man in his 50s who had sustained a spinal cord injury in a car accident more than 10 years before enrollment, resulting in severely impaired bimanual motor function and loss of effective grasping, holding, and pinching abilities. Limited right arm function was preserved, allowing him to raise his right forearm within a constrained range of motion. Preoperative neurological assessment showed complete C4 spinal cord injury (AIS-A; Table S1, Fig. S1). Patient 02 (P02) was a man in his 40s who had sustained a spinal cord injury 3 years before enrollment. He exhibited a similar pattern of upper-limb motor impairment, although his residual motor function was slightly better than that of P01. Preoperative neurological assessment showed incomplete C7 spinal cord injury (AIS-B; Table S2, Fig. S1). Informed consent was obtained from both participants before enrollment. In addition, consent for publication of materials depicting the participants was obtained.

For P01, muscle tone before surgery was rated as grade 2–3 on the Modified Ashworth Scale, indicating increased tone that affected passive movement within the range of motion. P01 also reported taking Baclofen (10 mg/day) and Pregabalin (75 mg/day) to relieve spasms at the sixth postoperative month; this symptom had persisted since the injury and was considered unrelated to the BCI implant.

##### S1.02 – Scalp EEG Motor Imagery Test

The EEG data of the patients were recorded using a 20-channel 10-20 EEG system cap (Pony, Neuracle). To eliminate interference from electromyographic (EMG) signals, only 12 electrodes located at the center area were selected for the study (F3, Fz, F4, C3, Cz, C4, P3, Pz, P4, O1, Oz,

O2), with CPz and AFz chosen as the reference and ground, respectively. The sampling rate was set to 1000 Hz.

The EEG motor imagery assessment paradigm involves both the left and right hands, with each round consisting of 15 trials. Each trial includes 5 seconds of movement, feedback (10 seconds for success, 2 seconds for failure), and 10 seconds of rest. Two rounds will be performed for each hand. At the beginning of the experiment, a cross will appear in the center of the screen for 10 seconds, during which the participant is instructed to focus on the cross and remain calm. After this period, the actual task begins. When the movement phase starts, an arrow pointing left or right will appear in the center of the screen, indicating left-hand or right-hand movement, respectively. A text prompt will also appear above the screen saying, "Imagine left-hand/right-hand movement." After 5 seconds, feedback is provided based on the system's evaluation of the participant's motor imagery EEG during that time. If successful, a celebratory animation will play, and a pneumatic hand will perform a single grasp and release action, lasting 10 seconds. If unsuccessful, a 2-second encouragement animation will be shown. Following this, there will be a 10-second rest period, during which an image of a teacup will appear on the screen with the text "Rest, relax, stay calm," and the participant will be asked to remain relaxed. Throughout the experiment, the participant is required to sit quietly in a wheelchair, focus on the screen, and wear the pneumatic hand device. When the corresponding prompt appears on the screen, the participant begins the associated task.

#### S1.03 – Multi-modal MRI-based Motor Localization

Before the implantation surgery, the subject underwent a head MRI on a 3.0T scanner (Siemens, Prisma) to locate brain areas associated with hand sensory and motor functions. Functional MRI (fMRI) was acquired using a gradient echo planar imaging (EPI) sequence with the following parameters: matrix size 100×100, resolution 2×2×2 $mm^3$, repetition time (TR) 2 s, echo time (TE) 30 ms. The scan covered the entire brain with 72 slices. T1-weighted MRI structural images were acquired using a magnetization-prepared rapid gradient-echo (MPRAGE) sequence with parameters: matrix size 256×256, resolution 1×1×1 $mm^3$, and 208 slices.

The fMRI paradigm was used to locate the subject's sensorimotor areas related to hand movement. The paradigm consisted of two parts. The first part involved passive movement, where two experimenters assisted the subject with passive fist clenching on the left and right sides at a frequency of approximately 0.5 Hz. This was designed to test whether sensory information could be transmitted to the cortex, indicating the condition of the subject's ascending pathways. The second part involved attempted movement, where the subject was asked to attempt to move their left or right hand in a familiar manner. This was used to locate the hand motor areas in the primary motor (M1) and primary sensory (S1) cortices. The block design included two alternating conditions: movement and rest. At the beginning of the movement condition, an arrow pointing left or right appeared on the screen with a text prompt to "start attempting left-hand/right-hand movement." At the beginning of the rest condition, an image of a teacup appeared with a text prompt to "rest, relax, stay calm," instructing the subject to remain relaxed during this period.

Each condition lasted 14 seconds (7 TRs, yielding 7 fMRI frames), repeated for 14 cycles. The left- and right-hand conditions alternated with a rest condition in between (Fig. S2).

#### S1.04 - Surgical Planning

Based on the results of the functional MRI activations, we developed a surgical implantation plan for the subject. Using the fMRI activation heatmaps combined with anatomical localization of the hand motor and sensory areas, we performed 3D modeling to select specific sites on the precentral and postcentral gyri for the right hand, where epidural cortical electrodes were placed to cover regions of significant fMRI activation (Fig. S2). The internal device was implanted into the flat skull behind the ear, ensuring sufficient bone thickness, avoiding the temporalis muscle, and facilitating wireless powering.

To effectively integrate the electrode positioning from the surgical plan with the surgical navigation system, an image registration method was developed to address compatibility issues between the 3D model and the navigation system. The cortex was segmented using FreeSurfer[1], and an individualized cortical model of the patient's brain was constructed. The electrode placement on the cortical surface was designed in Blender to cover the functional areas of the precentral and postcentral gyri. To import the positioning results into the surgical navigation system, we wrote the electrode locations into the patient's original T1 DICOM data. This process involved coordinate transformation between FreeSurfer's Surface RAS reference frame and the image's original RAS reference frame (Equation 1).

$$X' = AX \qquad 1$$

where $X$ represents the homogeneous coordinates of the electrode points in the Surface RAS reference frame$(x, y, z, 1)$, $X'$ represents the new coordinates in the original RAS reference frame $(x', y', z')$, and $A$ is the transformation matrix $(3 \times 4)$ converting from the FreeSurfer's Surface RAS coordinate system to the DICOM image's scanner RAS coordinate system. An optimized KD-tree nearest neighbor algorithm was used to efficiently map these coordinates to the pixel points in the original DICOM images. In this setup, all pixels within a 1mm radius were marked with a value significantly higher than the maximum MRI intensity (e.g., 5000). The modified DICOM files were then imported into the navigation system, allowing the surgeon to directly determine the electrode positions during surgery. The final electrode implantation sites, as shown in Fig. 1d, matched the planned locations and the areas of optimal fMRI activation.

#### S1.05 - Brain-Computer Interface Training and Evaluation Paradigms

To calibrate, update, and evaluate the brain-computer interface (BCI) system for hand motor control, structured calibration and daily-session paradigms were implemented throughout training and follow-up (Fig. S9). In both paradigms, the participant performed alternating blocks of attempted left- or right-hand movement and rest, each lasting 8 seconds. At the beginning of each attempted-movement block, a visual cue indicated the instructed hand, and the participant was asked to continuously attempt the corresponding hand movement during the task period. Rest blocks provided a baseline condition and separated adjacent movement-attempt trials.

The calibration session was used for personalized model initialization and early BCI testing. Each calibration session consisted of 30 rounds in total. In the first 15 rounds, the participant received sham feedback with an apparent accuracy of 85% to facilitate task engagement and adaptation to the training procedure. In the second 15 rounds, feedback was generated using the model calibrated from the first round of data, thereby providing true online feedback. During calibration, successful detection of attempted hand movement triggered actuation of the pneumatic glove, forming a closed-loop training condition. Data from the calibration session were used to initialize and update the motor-control decoding model.

After model establishment, daily sessions were performed to evaluate decoder performance and signal stability over time. The daily-session paradigm used the same alternating 8-second attempted-movement and rest structure but consisted of 10 rounds. In contrast to the calibration session, glove actuation in the daily session was triggered at task onset rather than by online detection of attempted hand movement, allowing standardized assessment across sessions while reducing variability introduced by decoder-dependent feedback.

In addition to these structured motor-control paradigms, functional BCI performance was assessed using a free-grasp task. In this task, the participant was instructed to use the BCI-controlled pneumatic glove to grasp a standard cylindrical object placed at the center of a nine-grid board and move it to one of eight peripheral target positions specified by the experimenter. This task was used to assess whether the participant could effectively use the BCI system to accomplish goal-directed object transfer under externally assigned target conditions. Together, these paradigms enabled model calibration, longitudinal performance evaluation, and functional assessment of BCI-assisted hand use.

#### S1.06 – Object-transfer Task and Assessment Time Points

To further quantify functional BCI-assisted grasp performance, an object-transfer task was performed in selected evaluation sessions. In this task, the participant used the pneumatic glove to grasp a standard cylindrical object placed at the center of a nine-grid board and move it to one of eight peripheral target positions specified by the experimenter, consistent with the free-grasp paradigm shown in Fig. S9b. A trial was counted as successful if the object was transferred from the center to the designated target position within 10 s. For each session, the average completion time of successful trials and the ratio of successful trials to total trials were calculated. The corresponding session-level results are summarized in Tables S4–S6.

Object-transfer testing was performed at defined postoperative time points during longitudinal follow-up. For P01, formal object-transfer testing was performed during postoperative days 180–184, and both BCI-assisted and unaided conditions were evaluated, allowing within-participant comparison of assistive benefit (Tables S4 and S5). For P02, formal object-transfer testing was performed on postoperative day 372, and only the BCI-assisted condition was evaluated (Table S6). The overall longitudinal training period for each participant is shown in Fig. S18.

Unaided object-transfer testing was not repeated in P02 because he had already regained voluntary grasping ability by the time of formal evaluation, and the purpose of this assessment was to quantify BCI-assisted task performance during follow-up rather than to re-test recovered bare-hand grasping ability.

#### S1.07 – Deep Learning based Hand Trajectory Tracing

To accurately capture hand movement trajectories during the free grasping process, a USB webcam was positioned directly above the experimental table to record the hand movements from a top-down perspective (Fig. S17). A serial port synchronization signal and LED light were used to synchronize the video recording with the ECoG data. For the video data of both pneumatic hand grasping and bare hand grasping, we finetuned a hand keypoint detection model to analyze the hand movement trajectories. The main structure of the model utilizes a high-resolution convolutional network (HRNet)[2] pre-trained on the OneHand10k dataset. The detection algorithm employs distribution-aware coordinate representation (DARK)[3] based on heatmaps to detect four key points of the pneumatic hand or the bare hand: the index fingertip, thumb tip, the base of the thumb (thenar), and the wrist. For the patient's grasping data, the coordinates of the thenar key point were extracted as the hand movement trajectory. The start time of each trial was defined as the moment the hand contacted the central cylindrical object, and the end time was defined as the moment of grasping failure or successfully reaching the target position and releasing the object. The model was then fine-tuned based on the OneHand10K pre-trained model. The pneumatic hand model training data included 1,295 annotated images with a resolution of 960x540, with 80% of the data used for training and 20% for testing. The keypoint recognition model achieved an AUC of 0.89. The bare hand detection model training data included 1,118 annotated images with a resolution of 960x540, with 80% of the data used for training and 20% for testing. The model achieved an AUC of 0.85.

### S2 – BCI Decoding

#### S2.01 - Geometry-Enhanced Multi-Spectral (GEMS) Decoder and Comparison Methods

The spatio-spectral information in epidural ECoG (eECoG) signals can be described using a covariance matrix. Attempted hand movement typically modulates eECoG activity across multiple frequency bands. By constructing a multi-band joint covariance matrix (Equation 3), we can effectively extract the spatio-spectral patterns of the EEG:

$$C_f = \begin{pmatrix} Z_{f_1}Z_{f_1}^T & \cdots & Z_{f_1}Z_{f_F}^T \\ \vdots & \ddots & \vdots \\ Z_{f_F}Z_{f_1}^T & \cdots & Z_{f_F}Z_{f_F}^T \end{pmatrix} \quad 2$$

where $Z_{f_i}, i = 1, \dots, F$ represents the channel signals filtered by different frequency bands, and F is the number of selected frequency bands.

The Affine Invariant Riemannian Metric (AIRM, Equation 4)[4] can measure the distance between covariance matrices:

$$\delta_r(C_1, C_2) = \left\| C_1^{-\frac{1}{2}} C_2 C_1^{-\frac{1}{2}} \right\|_F = \left( \sum_{i=1}^{n} \log^2 \lambda_i \right)^{\frac{1}{2}} \qquad 3$$

where $\|\cdot\|_F$ represents the Frobenius norm, and $\lambda_i$ are the eigenvalues of the matrix $C_1^{-1/2} C_2 C_1^{-1/2}$.

The AIRM metric has the following property: for any two samples from the source space that form covariance matrices $S_A$ and $S_B$, mapping through a propagation matrix $L$ (where $L$ is invertible) results in sensor space covariance matrices $X_A = LS_AL^T$, $X_B = LS_BL^T$. It can be proven that:

$$\delta_r(X_A, X_B) = \delta_r(S_A, S_B) \qquad 4$$

This indicates that the separability in the source space and the sensor space is identical. Similarly, over long-term use, physiological changes may alter the propagation matrix $L'$, but the AIRM distance measure ensures that separability remains unchanged. In contrast, BCI decoding algorithms using spatial filters may fail due to changes in the propagation matrix, causing spatial filter templates to become ineffective. Therefore, BCI decoding algorithms based on the AIRM metric possesses robustness in the sensor space and long-term stability.

To select the most effective frequency bands for decoding, we evaluated the separability of different frequency bands for rest and attempted right-hand movement using a Riemannian-based class distinctiveness method. A Fisher-type class separability metric under the Riemannian metric was adopted, in which class distinctiveness was quantified as the ratio of the inter-class distance to the average within-class dispersion (Equation 5)[5]:

$$D(A, B) = \frac{\delta_r\left(\overline{C^A}, \overline{C^B}\right)}{\frac{1}{2}(\sigma_{C^A} + \sigma_{C^B})} \qquad 5$$

where $\overline{C}$ represents the mean covariance under the Riemannian metric, and $\sigma_C$ represents the standard deviation of the covariance under the Riemannian metric, $\sigma_C = \frac{1}{n-1} \sum_{i=1}^{n} \delta_r\left(C_i, \overline{C}\right)$. The superscripts $C^{(A/B)}$ indicate the corresponding classes.

Subsequently, a classifier based on AIRM can be constructed by embedding the data into the tangent space[6]. For the manifold $M$ constituted by the aforementioned eECoG feature covariance matrices, the Fréchet mean point $X_{ref}$ of the manifold can be found (Equation 6):

$$X_{ref} = \underset{X_{ref}}{\operatorname{argmin}} \sum_{i=1}^{N} \delta^2 \left(X_i, X_{ref}\right) \quad 6$$

Based on this point, the tangent space $T_{X_{ref}}M$ can be constructed, and the data points $X_i$ can be embedded into this space to obtain the projected points $S_i$ (Equation 7), where logm denotes the matrix logarithm operation.

$$S_i = \log_{X_{ref}}(X_i) = X_{ref}^{-\frac{1}{2}} \operatorname{logm}\left(X_{ref}^{-\frac{1}{2}} X_i X_{ref}^{-\frac{1}{2}}\right) X_{ref}^{-\frac{1}{2}} \quad 7$$

In our study, the epidural intracranial EEG of patients exhibits a typical power increase at high frequencies (e.g., 55-95 Hz) and a typical power decrease at low frequencies (15-30 Hz and 35-50 Hz). Therefore, we designed a geometry-enhanced multi-spectral (GEMS) algorithm. Due to the physiological differences between low-frequency and high-frequency oscillatory activities, low-frequency activities were filtered at 15-30 Hz and 35-50 Hz, while high-frequency activities were band-pass filtered at 55-95 Hz and then enveloped (Fig. S3). Subsequently, a large covariance matrix of the feature signals was constructed, which was whitened to retain 99% of the variance dimension components to reduce the dimensionality. The samples were then embedded into the tangent space constituted by the Fréchet mean point, and finally sent to a logistic regression classifier to build the decoder for BCI calibration.

Comparison decoders. We benchmarked GEMS against a linear model[7,8]. Signals were filtered into nine bands spanning 0–150 Hz and downsampled to 10 Hz. Features from 7 channels, 9 bands and 5 time points formed a 315-dimensional vector for logistic regression.

The common spatial pattern (CSP) baseline[9,10] constructed covariance matrices using the same frequency bands as GEMS and retained 16 spatial filters. Their energy features were classified by logistic regression. For both baselines, regularization was selected by 10-fold cross-validation. Monthly decoding performance is shown in Fig. S7.

#### S2.02 - Hidden Markov Model for Temporal Regularization

Considering that the natural grasping process is inherently continuous, with each state at a given time point being dependent on the preceding and following states, leveraging this dependency can significantly enhance the reliability of maintaining a grasp. The hidden Markov model (HMM), by constructing a multi-order stochastic process, links the probability of the current hidden state with the observed data and the probability of the hidden state at the previous time point. This can be used to describe the natural grasping process. We define the hidden states as either resting or grasping. In our constructed HMM algorithm, the emission probability $p(s_k|z_t)$, which is proportional to the likelihood $p(z_t|s_k)$, is derived from the Riemannian geometric classification model obtained through supervised training, while the transition probability matrix is semi-supervised, estimated using the forward-backward algorithm based on data from the patient's free-grasping training sessions. The first-order Hidden Markov Model can predict the probability of each hidden state at the current step based on the probability of each hidden state at the previous step (Equation 8):

$$p(s_{k,t}) = p(s_k|z_t)\sum_i p\left(s_{k,t}|s_{i,t-1}\right)p(s_{i,t-1}) \quad 8$$

where $z_t$ is the observed data at time $t$, and $s_k$ is state $k$. We set a threshold $p_{th}$, and when the likelihood probability of the patient's new state exceeds this threshold, the model switches states. This threshold is estimated based on offline validation results and is set to 0.8 for the patient.

Additionally, we apply an extra step of first-order filtering to smooth the probabilities output by the HMM (Equation 9). The parameter $\eta$ is adjusted based on offline validation results and is set to 0.7 for the patient.

$$\hat{p}_{curr} = \eta p_{last} + (1-\eta)p_{curr} \quad 9$$

where $p_{last}$ is the hidden state probability from the previous step, $p_{curr}$ is the current hidden state probability inferred from Equation , and $\hat{p}_{curr}$ is the final estimated hidden state probability.

#### S2.03 - Model Training and Evaluation

The model is trained based on two sets of calibration data. Since high gamma (HG) activity decays over time during attempted grasping, we use data from the 0.5-3 seconds after the onset of attempted grasping and 4-7 seconds of resting state as the training data. The data are segmented at 0.5-second intervals for model fitting. One set of data is used for model training, utilizing 10-fold cross-validation to select the optimal regularization parameters for the classifier. After validation, a final model is trained using both sets of calibration data. After fitting the classification model, we fit the HMM state transition matrix with Baum–Welch expectation-maximization (EM) algorithm using the patient's own recordings from uncued (free) grasping sessions, in which the participant performs voluntary grasp–release movements without timed cues. Validation is conducted using a different set of data, evaluating both segment metrics and event metrics.

During model calibration, appropriate evaluation criteria are required to assess decoder performance. Because the goal is to decode natural grasping, which unfolds as a continuous temporal process, both slice-level and event-level performance should be evaluated.

**Slice-level metrics.** The decoder first classifies short data segments, and slice-level performance is quantified using classification accuracy (Fig. 3e-f) or area under ROC curve (AUC, Fig. S7).

**Event-level metrics.** To evaluate detection performance in continuous time, we further defined event-based metrics. A grasping event was counted as correctly detected if at least one decoder output occurred within a window extending from 0.5 s before grasp onset to 2 s after onset; otherwise, it was counted as a false negative. If multiple detections were produced for the same grasping event, the additional detections were counted as false positives. Event-level precision, recall, and F1 score were then calculated accordingly. Because grasping events were sparse in the continuous recordings, chance performance for event detection was substantially lower than that of a balanced binary classification task. Notably, the chance level of the event-level F1 score is not a universal constant, but depends on the temporal density of true events, the matching window used to define correct detections, and the false-alarm rate of the detector. Under the temporal design of our calibration task, the chance-level event F1 was 0.15, reflecting the difficulty of continuous grasp detection compared with simple slice-wise binary classification.

### S3 - Offline Analysis of EEG/ECoG data

#### S3.01 - Comparison of Three Types of Brain Electrophysiology Recordings

To compare the differences between epidural electrocorticography (eECoG) and subdural electrocorticography (sECoG), we utilized data from the publicly available Kai Miller dataset[11]. Since the cortical activity patterns during attempted movement in patients with paralysis are more similar to those during actual movement in healthy individuals[12], we selected real hand movement data from 10 patients whose electrodes covered the hand area. The cortical electrodes in the Kai Miller dataset had a spacing of 10 mm and a contact diameter of 2.3 mm. In our study, we used electrodes with a spacing of 8 mm and a contact diameter of 3.2 mm, which are comparable in size. The EEG electrode spacing in the patients' EEG caps was approximately 4 cm. To ensure consistent results, all three types of brain electrical activity were processed using the common average re-reference method. Additionally, to study the spatial resolution differences between different brain recordings, we introduced scalp EEG motor imagery data from the BCI2000 dataset for comparison[13].

We quantified frequency-domain modulation by calculating the relative power change in eECoG, sECoG, and EEG during 0–2.5 s of movement with respect to 0–2.5 s of rest (Equation 10). This analysis allowed us to compare the magnitude of task-related spectral modulation across frequency bands and recording modalities.

$$Power\ Change = \frac{P_{task} - P_{rest}}{P_{rest}} \tag{10}$$

Where $P_{task}$ refers to the power of the brain electrical activity in the frequency band during movement, and $P_{rest}$ refers to the power in the resting state. Power spectral density (PSD) was calculated using resting state data to compare the spectral energy differences of the different brain electrical activities (Fig. 2b).

To qualitatively compare the spatial resolution differences, we calculated the relationship between the distance between electrode pairs and the correlation coefficient of the electrode signals (Fig.

2c). To maintain consistency with the number of subjects and tasks selected in the eECoG dataset, we selected all electrode pairs from the first 10 subjects in the BCI2000 dataset for real left- and right-hand movements. All eECoG signals were filtered with a 1 Hz high-pass filter, using the original reference electrodes (EEGs were referenced to mastoids, sECoGs and eECoGs were referenced to the scalp).

#### S3.02 - Demixed Principal Component Analysis of Daily Spatial Covariance Patterns

To track the longitudinal evolution of condition-specific neural activity patterns in the implanted ECoG array, we developed a visualization pipeline that combines Riemannian geometry on the manifold of symmetric positive-definite (SPD) matrices with demixed principal component analysis (dPCA). This approach yields a low-dimensional embedding in which each recording day is represented as a point, colour-coded by date and motor condition, enabling visual inspection of how neural covariance structure evolves over months of BCI training.

##### Spatial Covariance Estimation

For each recording day, ECoG signals are bandpass-filtered into pre-defined frequency bands (e.g., β: 15–35 Hz; high-γ: 55–95 Hz). For high-frequency bands, the analytic amplitude envelope is additionally extracted via the Hilbert transform. Within each band, time-locked epochs are cut around paired motor-task events (e.g., grasp onset and rest onset), and the spatial covariance matrix of each single-trial epoch is estimated using a Ledoit–Wolf shrinkage estimator to regularize the sample covariance under limited trial counts.

Trial-level covariance matrices within a day are then averaged on the Riemannian manifold of SPD matrices. The Riemannian (Fréchet) mean is defined in Equation 6. The procedure is performed independently for each condition (grasp, rest) and each frequency band, producing per-day summary covariance matrices $\bar{\boldsymbol{S}}_{d,b}^{(\mathrm{grasp})}$ and $\bar{\boldsymbol{S}}_{d,b}^{(\mathrm{rest})}$ for day $d$ and band $b$.

##### Tangent-Space Projection

Because SPD matrices reside on a non-Euclidean manifold, standard linear dimensionality reduction cannot be applied directly. We therefore map all daily covariance matrices into a common tangent space using the logarithmic map at a reference point (Equation 7), yielding a feature vector $\boldsymbol{v}_i \in \mathbb{R}^{C(C+1)/2}$. In this tangent space, Euclidean operations such as PCA yield geometrically faithful approximations to geodesic computations on the manifold.

##### Demixed Principal Component Analysis

Standard PCA applied to the tangent-space vectors would find axes of maximal total variance, mixing variance due to condition differences with variance due to day-to-day drift and their interaction. Demixed PCA (dPCA)[14,15] instead decomposes the data into interpretable marginal components before extracting principal axes, yielding projections that can be attributed to specific experimental factors.

#### Marginal decomposition

The tangent-space feature vectors are arranged into a three-way tensor $\mathcal{T} \in \mathbb{R}^{F \times Q \times D}$, where $F = C(C+1)/2$ is the tangent-space feature dimensionality, $Q = 2$ is the number of conditions (grasp, rest), and $D$ is the number of recording days. This tensor is decomposed according to an ANOVA-like additive model:

$$\mathcal{T}_{fqd} = \bar{\mu}_f + \phi_{fq}^{(cond)} + \phi_{fd}^{(day)} + \phi_{fqd}^{(int)} \tag{11}$$

where:

$\bar{\mu}_f = \frac{1}{QD} \sum_{q,d} \mathcal{T}_{fqd}$ is the grand mean over conditions and days;

$\phi_{fq}^{(\mathrm{cond})} = \frac{1}{D} \sum_d \mathcal{T}_{fqd} - \bar{\mu}_f$ is the condition marginal, capturing the average difference between grasp and rest covariance geometry;

$\phi_{fd}^{(\mathrm{day})} = \frac{1}{Q} \sum_q \mathcal{T}_{fqd} - \bar{\mu}_f$ is the day marginal, capturing condition-independent longitudinal drift;

$\phi_{fqd}^{(\mathrm{int})} = \mathcal{T}_{fqd} - \bar{\mu}_f - \phi_{fq}^{(\mathrm{cond})} - \phi_{fd}^{(\mathrm{day})}$ is the interaction residual, reflecting day-specific deviations in condition separability.

#### Axis selection and orthogonalization

For each marginal component, the corresponding sub-tensor is unfolded into a two-dimensional matrix by collapsing the condition and day dimensions into a single sample axis, and singular value decomposition (SVD) is applied to extract the leading principal direction(s). Specifically, for each marginal $m \in \{\mathrm{cond}, \mathrm{day}, \mathrm{int}\}$, the unfolded matrix $\boldsymbol{M}^{(m)} \in \mathbb{R}^{(Q \cdot D) \times F}$ is mean-centred and decomposed as $\boldsymbol{M}^{(m)} = \boldsymbol{U\Sigma V}^{\top}$. The first right singular vector of each marginal, along with leading axes from the global (un-decomposed) centred tensor, form a candidate pool.

The final projection axes are constructed by a greedy sequential Gram–Schmidt procedure: candidate axes are considered in order of marginal priority (condition, day, interaction, global), and each is orthogonalised against the previously selected axes. A candidate is accepted if its residual norm after orthogonalisation exceeds machine precision; otherwise it is discarded. This yields an orthonormal set of two axes spanning the directions of greatest condition-related and time-related variance in covariance geometry. Each axis inherits a label indicating which marginal it primarily represents (e.g., “dPC (condition)”, “dPC (day)”).

##### Day-Level Cross-Validation

To guard against overfitting the low-dimensional projection to the specific set of recording days, we employ $K$-fold cross-validation at the day level. Days are randomly partitioned into $K$ folds (default $K = 10$). In each fold, the dPCA axes are fit exclusively on the training days; the held-out test days are then projected onto these axes. To prevent arbitrary sign flips across folds, each fold’s axes are aligned in sign to a reference set fit on the full dataset, by flipping axes whose inner product with the corresponding reference axis is negative.

This procedure ensures that the two-dimensional coordinates of every day are obtained from axes that were not optimized on that day’s data, providing an honest low-dimensional summary of the longitudinal covariance evolution.

### S4 - Electrophysiological and Neurological Assessment of Rehabilitation

#### S4.01 - SEP Measurement

The testing of somatosensory evoked potentials (SEP) follows the standards of the international federation of clinical neurophysiology (IFCN)[16]. The stimulation sites are at the wrist corresponding to the median, radial and ulnar nerves respectively. A 6×9 cm silver/silver chloride adhesive electrode is used. The positive electrode is placed distally on the trunk, the negative electrode proximally, with a 2 cm distance between them. The stimulation is a unidirectional pulse, with 2 Hz frequency 400 μs pulse width and amplitudes of 16 mA for the median and radial nerves, or 12 mA for the ulnar nerve. The stimulations and brain responses are synchronized based on NEO’s trigger system, which ensures sub-millisecond synchronization precision. This setup aims to stably evoke SEP while avoiding pain. The stimulation is repeated 160 times in each session. During stimulation, SEPs are recorded with epidural electrodes of NEO BCI system at a sampling rate of 1000 Hz. The signals are band-pass filtered (20-400 Hz), baselined according to the average of 80 ms data before stimulation onsets, and averaged across trials to obtain the SEP waveform. Monthly recordings are repeated at the same stimulation site and parameters to observe changes in the SEP waveform.

In SEP analysis, for early (15-35 ms[17]) and late (40-80 ms[18]) SEP components, the average absolute amplitude is extracted[19]. To minimize amplitude increases due to enhanced random phase oscillatory activity, the Bootstrap method is used. For each Bootstrap sample set, the average SEP response is extracted, and the average waveform amplitudes of early and late components are extracted. For the data from the first and the last month after implantation, permutation test is used to calculate the p-values that the last month’s amplitude is greater than that of first month (for P01 are M1 and M24, and for P02 are M1 and M12), and the FDR correction is applied to control the false discovery rate in multiple comparisons.

##### S4.02 - Neurological Scales Measurement

This study uses the international standards for neurological classification of spinal cord injury (ISNCSCI) scale[20] and the action research arm test (ARAT) scale[21] to assess patients' neurological and upper limb functional status, respectively. ISNCSCI evaluations were performed by two attending physicians, and any discrepancies were resolved by discussion to obtain a consensus result. The longitudinal assessment focused on changes in upper-limb motor scores and sensory scores across follow-up sessions. ARAT assessments were conducted by rehabilitation physicians and experimenters, and the testing process was video-recorded (Video 2). These assessments provided complementary measures of neurological status and functional hand performance throughout the rehabilitation period.

#### S5 – Modeling of BCI Training–Induced Functional Recovery

To investigate the dynamical mechanism of rehabilitation, we implemented a reduced corticospinal sensorimotor model consisting of four interacting populations: descending drive from motor cortex (C), peripheral afferent feedback (A), spinal interneurons (E), and α-motoneurons (M) (Fig. 5f) [22]. Cortical activity represented the intended motor drive, whereas afferent activity represented pneumatic glove-mediated sensory feedback. Both streams converged onto spinal interneuron and motoneuron populations, thereby closing a simplified sensorimotor loop. The corticospinal projection from C to E was initialized at weak strength to mimic spinal cord injury and was allowed to evolve through spike-timing-dependent plasticity (STDP)[23,24].

##### S5.01 - Neuron and synapse dynamics

The E and M populations were modeled as leaky integrate-and-fire neurons. For each neuron, the membrane potential v(t) and synaptic current I(t) evolved according to:

$$\frac{dv}{dt} = \frac{V_{\text{rest}} - v}{\tau_m} + \frac{I}{\tau_m} \tag{12}$$

$$\frac{dI}{dt} = -\frac{I}{\tau_{\text{syn}}} \tag{13}$$

A spike was emitted when $v > V_{\text{th}}$, after which the membrane potential was reset to $V_{\text{reset}}$ and held refractory for a period $t_{ref}$. In the simulations, $V_{\text{rest}} = -65\text{mV}$, $V_{\text{reset}} = -65\text{mV}$, $V_{\text{th}} = -50\text{mV}$, $\tau_m = 20\text{ms}$, $\tau_{\text{syn}} = 5\text{ms}$, and $t_{\text{ref}} = 2\text{ms}$. The numerical integration time step was 0.1 ms, and each simulation lasted 6 s.

Cortical and afferent drives were implemented as Poisson spike generators with externally specified time-dependent firing rates $rate_{\mathrm{C}(t)}$ and $rate_{\mathrm{A}(t)}$. In the baseline simulation, the cortical input window was defined on $[C_{\mathrm{on}}, C_{\mathrm{off}}] = [1.0s, 5.0s]$, whereas the afferent input window was defined on $[A_{\mathrm{on}}, A_{\mathrm{off}}] = [1.5s, 5.0s]$.

#### S5.02 - Connectivity and synaptic weights

Network connectivity was sparse and probabilistic. The connection probabilities were $p_{CE} = 0.3$, $p_{AE} = 0.9$, $p_{EM} = 0.04$, and $p_{AM} = 0.9$ for the projections C→E, A→E, E→M, and A→M, respectively. Fixed synaptic efficacies were used for all pathways except C→E: $w_{AE} = 1.80\mathrm{mV}$, $w_{EM} = 0.35\mathrm{mV}$, and $w_{AM} = 1.50\mathrm{mV}$. The corticospinal synapses C→E were initialized at $w_{CE,0} = 0.002\mathrm{mV}$, representing markedly weakened descending transmission after injury [25]. For all synapses, presynaptic spikes incremented postsynaptic current according to $I_{\mathrm{post}} \leftarrow I_{\mathrm{post}} + w$.

#### S5.03 - STDP rule for corticospinal plasticity

Plasticity was applied only to the corticospinal C→E synapses. Each such synapse carried a presynaptic trace $\mathrm{x_{pre}}$ (t) and a postsynaptic trace $\mathrm{x_{post}}$ (t), which decayed exponentially as:

$$\frac{d\mathrm{x_{pre}}}{dt} = -\frac{\mathrm{x_{pre}}}{\tau_{\mathrm{pre}}} \tag{14}$$

$$\frac{d\mathrm{x_{post}}}{dt} = -\frac{\mathrm{x_{post}}}{\tau_{\mathrm{post}}} \tag{15}$$

Both time constants were set to $\tau_{\mathrm{pre}} = \tau_{\mathrm{post}} = 20\mathrm{ms}$. At each presynaptic spike, the presynaptic trace was incremented by 1 and the synaptic weight was depressed according to:

$$\Delta w = A_{-} \cdot x_{post} \tag{16}$$

$$w \leftarrow \begin{cases} w_{\mathrm{min}}, if \;\; w - \Delta w < w_{\mathrm{min}} \\ w - \Delta w, if \;\; w_{\mathrm{min}} \leq w - \Delta w \leq w_{\mathrm{max}} \\ w_{\mathrm{max}}, if \;\; w - \Delta w > w_{\mathrm{max}} \end{cases}$$

At each postsynaptic spike, the postsynaptic trace was incremented by 1 and the synaptic weight was potentiated according to:

$$\Delta w = A_{+} \cdot x_{pre} \tag{17}$$

$$w \leftarrow \begin{cases} w_{\mathrm{min}}, if \;\; w + \Delta w < w_{\mathrm{min}} \\ w + \Delta w, if \;\; w_{\mathrm{min}} \leq w + \Delta w \leq w_{\mathrm{max}} \\ w_{\mathrm{max}}, if \;\; w + \Delta w > w_{\mathrm{max}} \end{cases}$$

The learning rates were $A_+ = 0.004$ mV and $A_- = 0.0045$ mV, with hard bounds $w_{\min} = 0$ and $w_{\max} = 0.5$ mV. This rule implements activity-dependent strengthening when cortical and spinal interneuron activity are temporally coordinated, and weakening when pre- and postsynaptic events are mismatched.

#### S5.04 - Readout of corticospinal recovery

To quantify the evolution of corticospinal coupling, synaptic weights from a subset of C→E synapses were recorded during the simulation. The mean corticospinal weight was computed as:

$$\bar{W}_{CE}(t) = \frac{1}{N_{\text{rec}}} \sum_i w_i(t) \quad (18)$$

where $N_{\text{rec}}$ denotes the number of recorded corticospinal synapses and $w_i(t)$ is the instantaneous weight of the i-th synapse. In the implementation, 20 representative synapses were monitored at 5-ms resolution, and the reported trajectory corresponded to the average across those synapses. This mean weight served as the principal model-level measure of corticospinal recovery.

#### S5.05 - Phenomenological mapping from corticospinal coupling to force and motor scores

To capture the nonlinear relationship between neural drive and macroscopic biomechanical output, we mapped the simulated synaptic weight to a normalized maximum voluntary force $F_{max}$ , using a logistic saturating function[26]:

$$F_{\max}(W_{CE}) = F_{\min} + \frac{F^*_{\max} - F_{\min}}{1 + \exp\left(-\frac{W_{CE} - W_{50}}{k_W}\right)} \quad (19)$$

Here $F_{min}$ denotes the minimum residual force, $F^*_{\max}$ denotes the maximal recoverable force available prior to rehabilitation. The transition is governed by $W_{50}$, the threshold synaptic weight required to achieve 50% of the maximum dynamic range, and $k_W$, which determines the steepness (gain) of the force recruitment curve

The parameters used were $F_{\min} = 0.05$, $F^*_{\max} = 1.0$, $W_{50} = 0.45$, and $k_W = 0.08$.

To bridge the gap between biomechanical force and standardized clinical assessments, we modeled the expected motor score based on $F_{max}$. We conceptualized the motor score as a battery of K distinct motor items, where the successful completion of the k item depends probabilistically on the patient's available force crossing a specific task-dependent threshold, $F_k$.

The probability $P_k$ of successfully completing item k is defined by a logistic function:

$$P_k = \frac{1}{1 + \exp\left(-\frac{F_{\max} - F_k}{\beta}\right)} \qquad 20$$

The expected motor score was computed as:

$$\mathbb{E}[\mathrm{MOTOR}](F_{\max}) = s_{\max} \sum_{k=1}^{K} P_k(F_{\max}) / \mathrm{K}s_{\max} \qquad 21$$

where K denotes the number of motor test items included, $s_{\max}$ is the maximum score per item, and β controls the smoothness of the force-to-performance transition. In the implementation, thresholds were linearly spaced from 0.15 to 0.65 and $\beta = 0.06$.

#### S5.06 - Training conditions and effects of feedback timing

Within this reduced model, passive glove-assisted training is represented by strong afferent drive in the absence of temporally coordinated cortical input, such that the A→E and A→M pathways activate spinal and motor populations without consistently engaging STDP at the corticospinal synapses. By contrast, BCI-mediated training is represented by temporally paired cortical activity and glove-evoked afferent feedback, which co-activate the C and E populations and thereby promote progressive strengthening of the C→E pathway through the STDP rule defined above. In the phenomenological readout, increases in corticospinal coupling were associated with higher normalized maximal force and higher expected motor performance.

To assess how feedback timing affects BCI-mediated plasticity within a single training trial, we varied only the onset delay $\Delta t$of afferent input $A$relative to cortical input $C$, while keeping the descending motor drive, the statistical properties of afferent input, and the plasticity parameters unchanged. Here, feedback onset delay denotes the time required for the brain–computer interface to decode motor intention and trigger the external device to generate sensory feedback.

The delay parameter $\Delta t$was swept from 0 to 4000 ms in increments of 200 ms. For each delay condition, 10 independent simulations were performed. In each repetition, new Poisson spike trains were generated for both $C$and $A$using the same rate definitions as in the baseline model, so as to reduce the incidental influence of any specific spike pattern on the STDP outcome. Each simulation corresponded to one complete grasp training trial. At the end of each trial, the mean synaptic weight change was computed for the C→E and A→E pathways as

$$\Delta W_X(\Delta t) = \bar{W}_{X,\mathrm{end}}(\Delta t) - \bar{W}_{X,\mathrm{start}}(\Delta t) \qquad 22$$

where $X \in \{CE, AE\}$, and $\bar{W}_{X,\mathrm{start}}$and $\bar{W}_{X,\mathrm{end}}$denote the mean synaptic weights at the beginning and end of the training trial, respectively. For each value of $\Delta t$, the mean and standard deviation across the 10 repetitions were then calculated.

To compare the magnitude of weight change across delay conditions, the resulting weight-change measure was further normalized by min–max scaling:

$$\Delta W_{\mathrm{norm}}(\Delta t) = \frac{\Delta W(\Delta t) - \Delta W_{\mathrm{min}}}{\Delta W_{\mathrm{max}} - \Delta W_{\mathrm{min}}} \quad 23$$

such that the maximum observed $\Delta W$was mapped to 1 and the minimum to 0. The model indicates that favorable weight updates are obtained when $\Delta t$ falls within a limited time range.

### Supplemental Figures

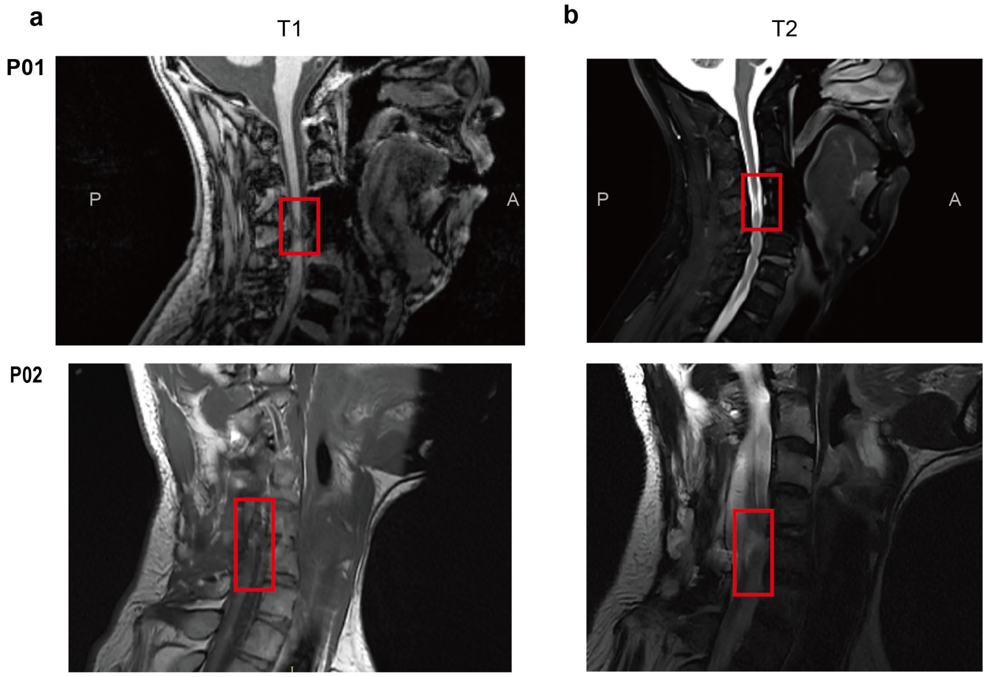

Figure S1 MRI scans of the patients' neck regions.

Sagittal cervical spine MRI from two patients (P01 and P02) is shown. a, T1-weighted structural images; b, T2-weighted structural images. The red boxes indicate the location of the spinal cord injury in each patient.

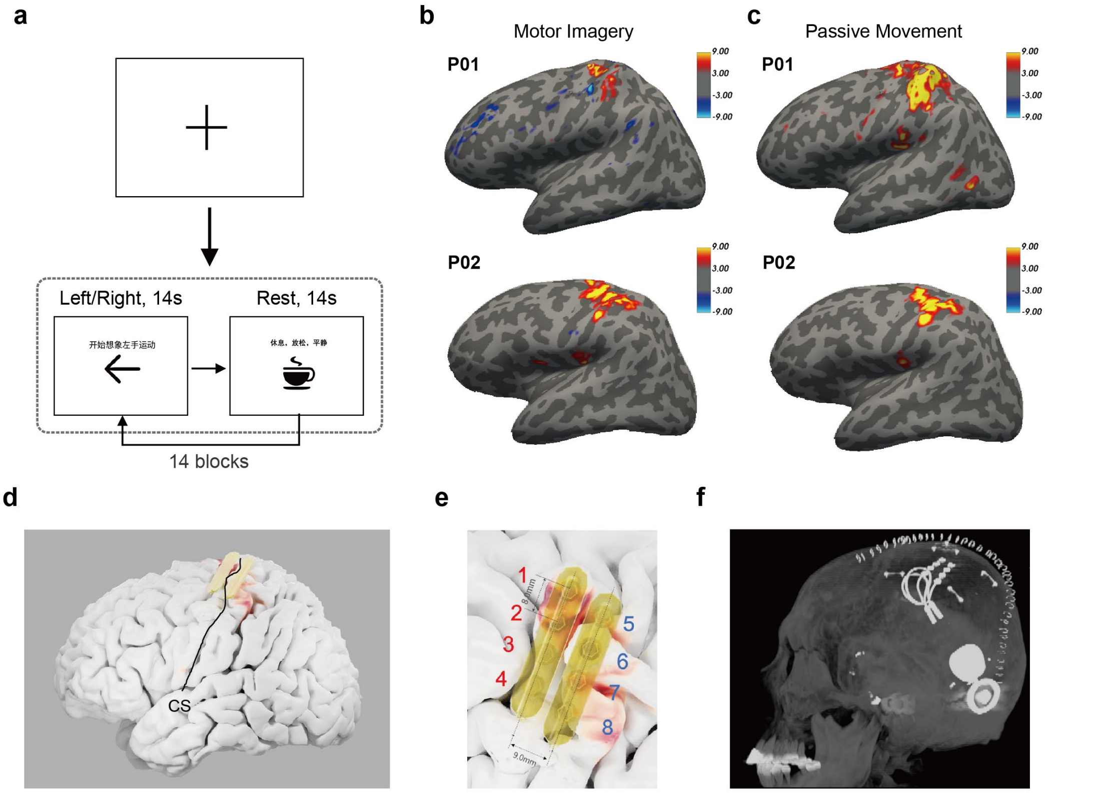

Figure S2 fMRI localization task, functional mapping, and surgical planning in two patients (P01 and P02).

a, Schematic of the fMRI localization task, with alternating rest and left- or right-hand movement conditions. b–c, Functional activation maps from P01 and P02 during attempted movement (b) and passive movement (c), with activation values expressed as −lg(p). d, Electrode planning based on the functional localization results. e, Enlarged view of the planned electrode arrangement; electrodes 1–4 were located on the precentral gyrus, whereas electrodes 5–8 were located on the postcentral gyrus. f, Sagittal postoperative CT image showing the actual electrode implantation positions. CS, central sulcus.

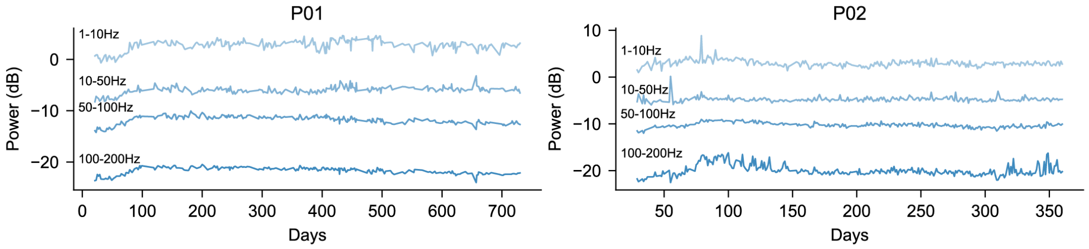

Figure S3 Longitudinal band-power changes in P01 and P02.

Time courses of ECoG power in four frequency bands (1–10 Hz, 10–50 Hz, 50–100 Hz, and 100–200 Hz) are shown for P01 (left) and P02 (right). Power is given in dB and plotted against days.

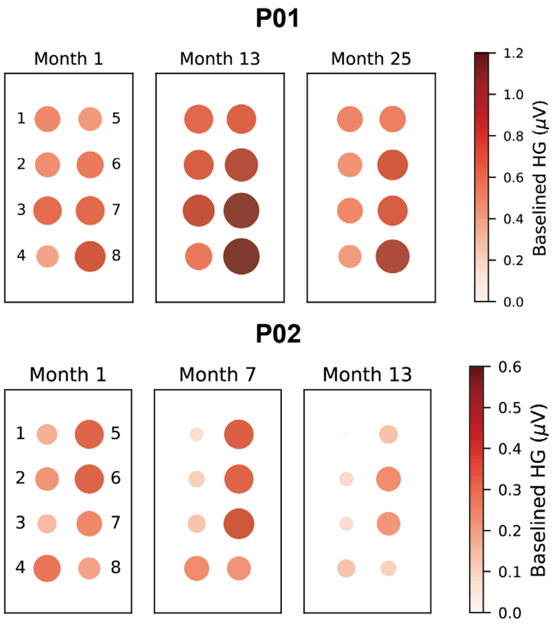

Figure S4 Spatial distribution of baseline high-gamma activity across months in the two patients.

Baseline high-gamma (HG) amplitude is shown for channels 1–8 in **P01** (top) and **P02** (bottom) at different recording months. Both circle size and color encode the amplitude of baseline HG activity, with darker and larger circles indicating higher values. Color bars denote amplitude in microvolts (μV).

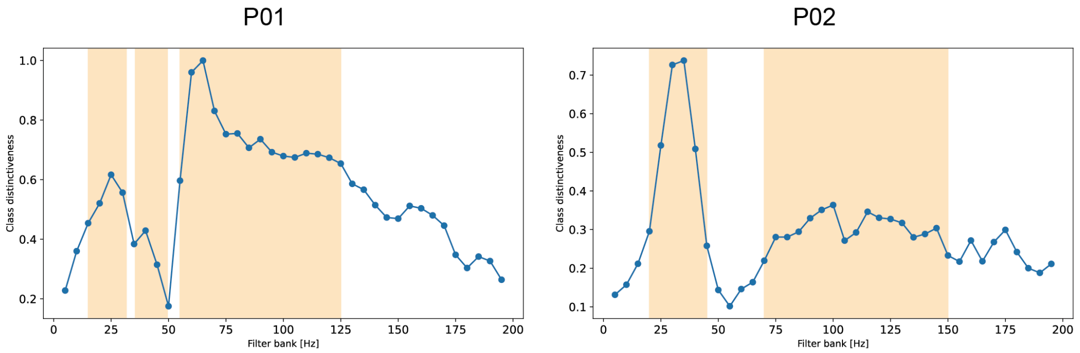

Figure S5 Frequency-band class distinctiveness in the two patients.

Class distinctiveness across filter banks is shown for P01 (left) and P02 (right). Yellow shaded regions denote the frequency bands used to train the final classifier. The spectral profiles differed between the two patients, indicating patient-specific distributions of discriminative information.

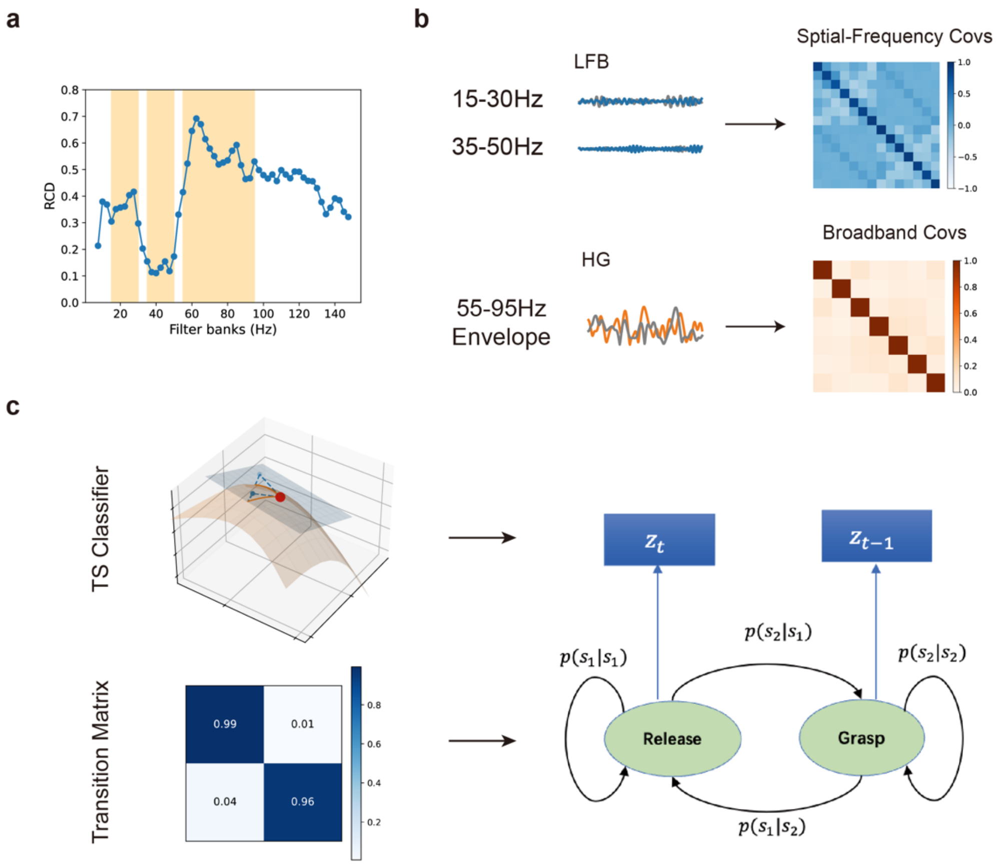

Figure S6 GEMS (Geometry-Enhanced Multi-Spectral) decoding pipeline.

a, Selection of informative frequency bands using Riemannian class distinctiveness (RCD) across filter banks. b, Extraction of covariance features from low-frequency band-pass activity and high-gamma envelope activity. c, HMM-based decoding framework for natural grasping, in which temporal dependencies between grasp states were modeled to enhance the stability and reliability of continuous decoding. The observation probabilities were provided by a trained Riemannian tangent-space classifier, and the transition matrix was fitted using patient behavioral data. TS classifier, tangent-space classifier.

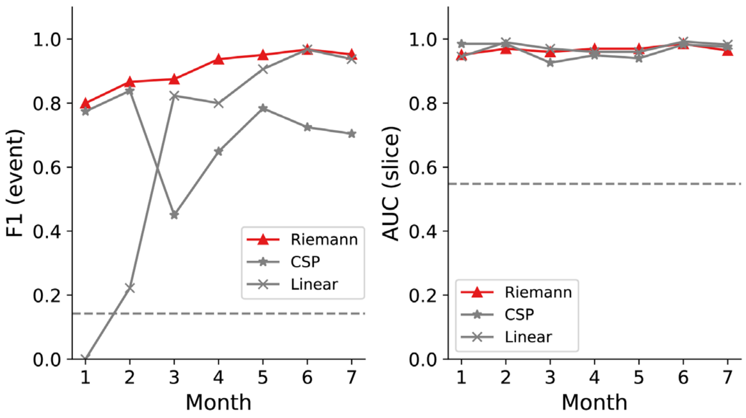

Figure S7 Longitudinal decoding performance in Patient 1 across seven months.

Left, event-level F1 score; right, slice-level AUC. Performance is shown for the Riemann, CSP, and linear methods at each monthly evaluation. The dashed horizontal lines indicate chance-level performance. Overall, the Riemann method showed consistently high performance across sessions, whereas CSP and the linear method exhibited greater variability over time.

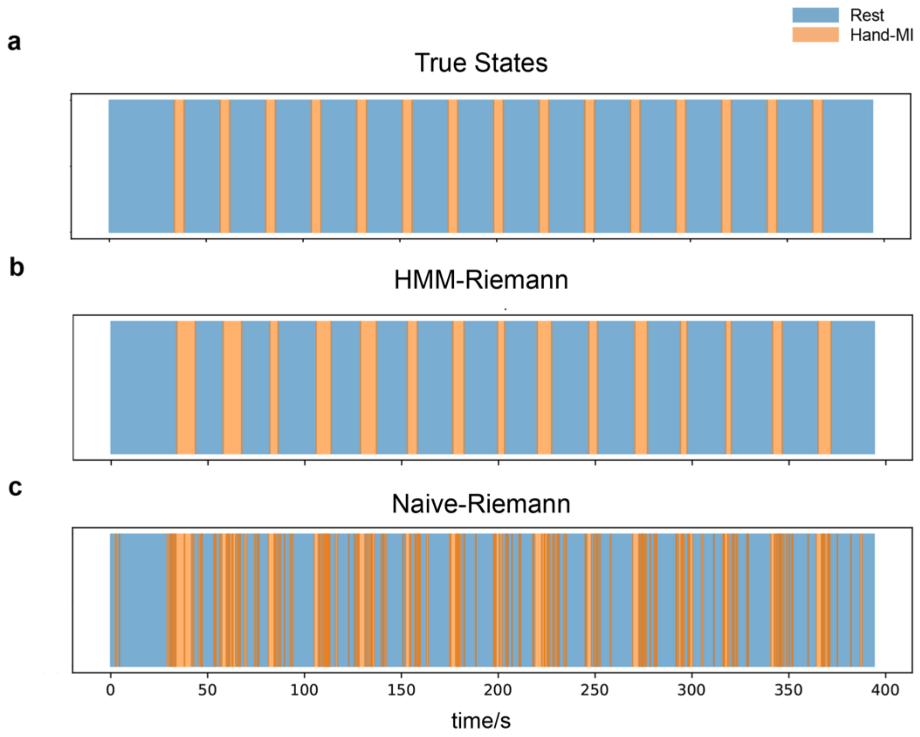

Figure S8 Comparison of continuous decoding with and without HMM modeling.

a, Ground-truth cues for attempted hand movement. b, Continuous decoding results generated by the HMM-Riemann method. c, Continuous decoding results generated by the Naïve-Riemann method without HMM, showing less stable state estimation. Hand-MA, hand movement attempt.

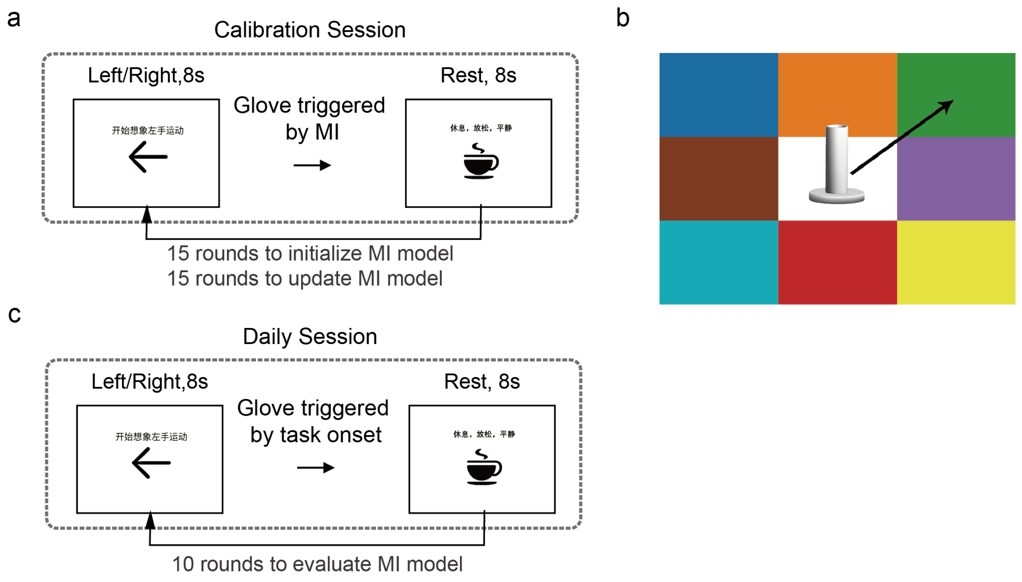

Figure S9 Calibration and training paradigms.

a, BCI calibration session. The paradigm consisted of alternating periods of attempted left- or right-hand movement and rest (8 s each). During the calibration session, the glove was triggered by detection of attempted hand movement. Fifteen rounds were used to initialize the motor-control decoding model, followed by 15 additional rounds to update the model. b, Free-grasp BCI task, in which the patient was instructed to move the standard cylinder from the central white grid to one of the eight colored peripheral grids specified by the experimenter. c, Daily session paradigm. The paradigm also consisted of alternating periods of attempted left- or right-hand movement and rest (8 s each), but the glove was triggered at task onset. Ten rounds were performed to evaluate the motor-control decoding model.

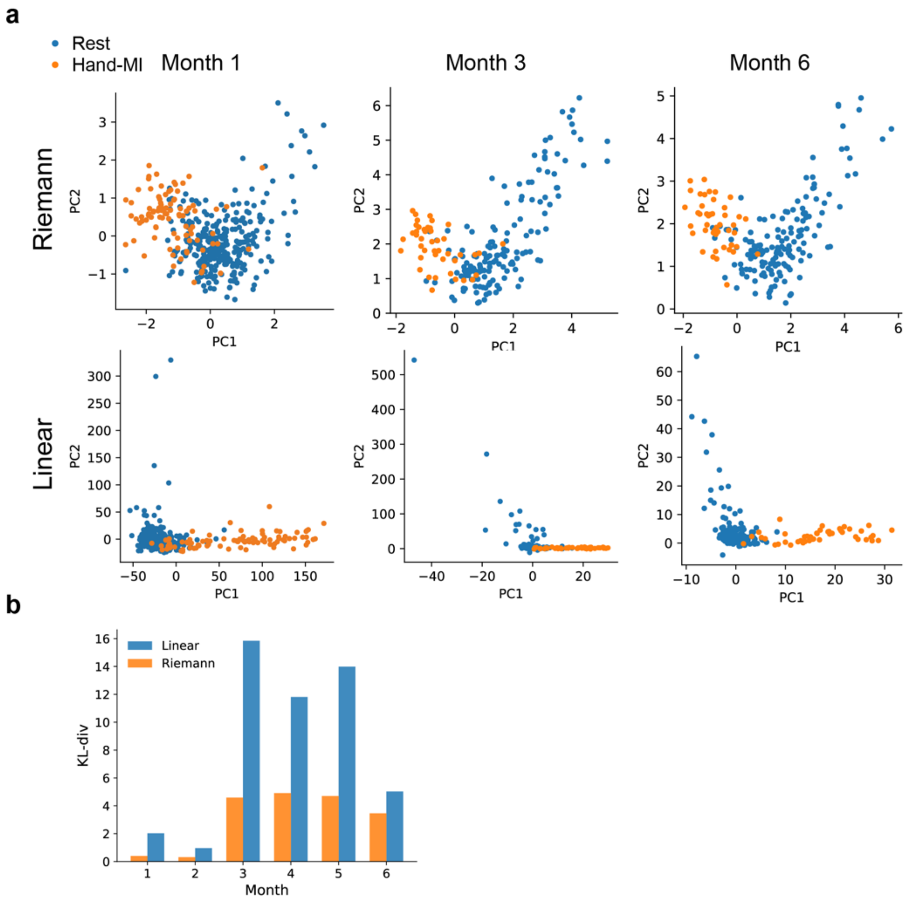

Figure S10 Longitudinal evolution of epidural ECoG feature distributions.

a, PCA projections of Riemannian features and linear spatio-temporal-spectral features across different months. Blue and orange points represent rest and attempted hand movement (Hand-MA), respectively. b, KL divergence from the training distribution across months for the two feature representations. Riemannian features exhibited smaller deviations from the training distribution than linear spatio-temporal-spectral features, suggesting superior stability during long-term use. KL-div, Kullback–Leibler divergence.

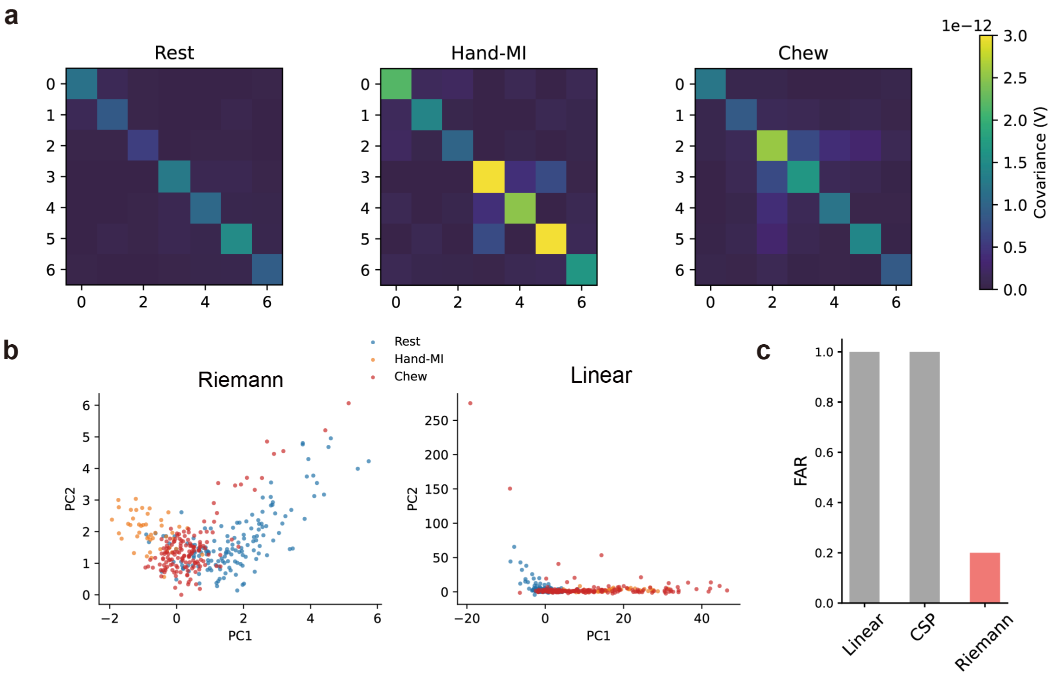

Figure S11 Characterization of EMG noise in different feature spaces.

a, Mean covariance patterns for rest, attempted hand movement (Hand-MA), and chewing EMG noise. b, Dimensionality-reduced representations of Riemannian features (left) and linear spatio-temporal-spectral features (right). In the Riemannian space, chewing EMG noise occupied an intermediate position between rest and Hand-MA, whereas in the linear feature space it largely overlapped with the Hand-MA distribution. c, Comparison of false acceptance rate (FAR) across decoding methods, demonstrating superior robustness of the Riemannian method relative to the linear and CSP methods.

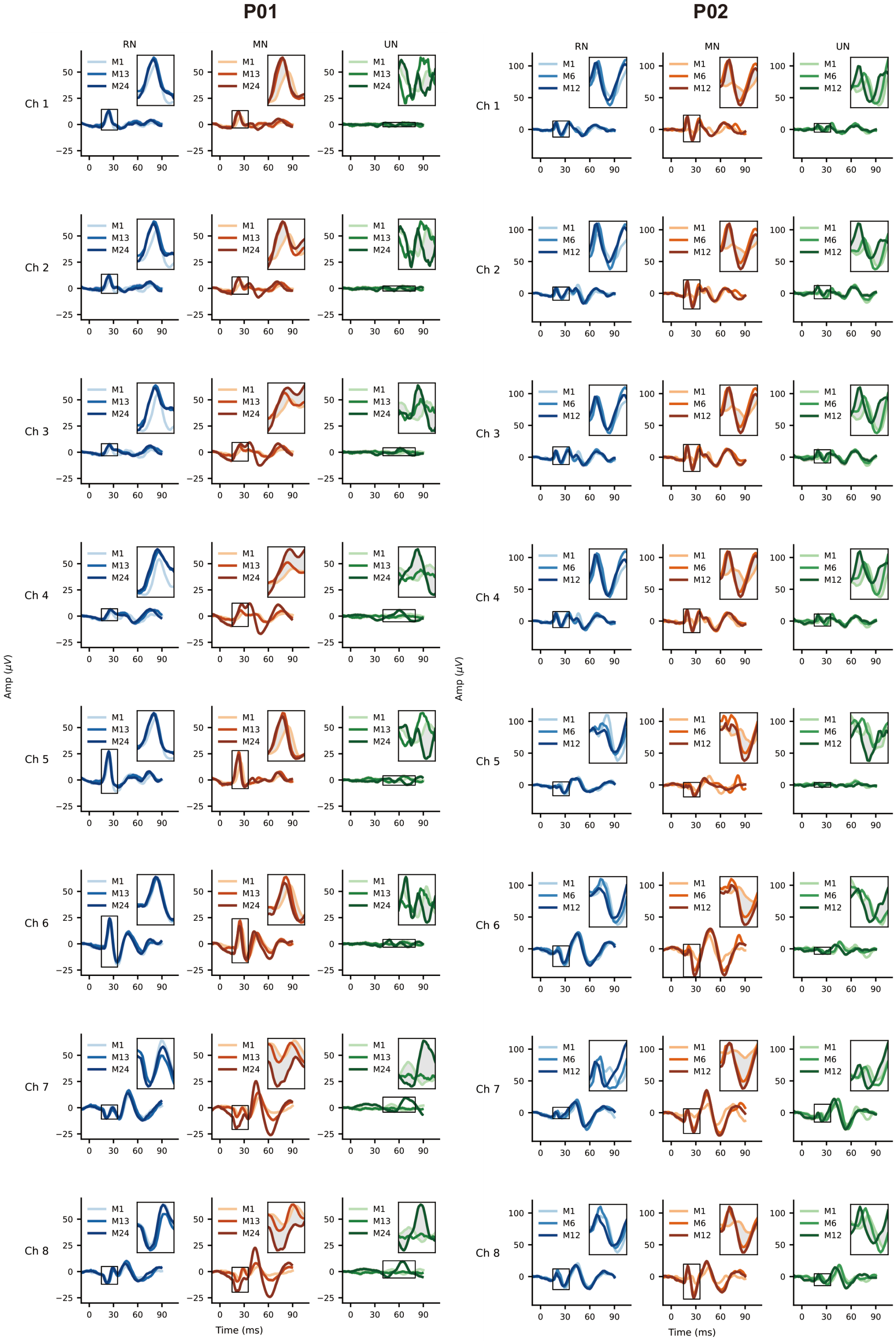

Figure S12 Longitudinal somatosensory evoked potentials in two patients.

SEP responses from P01 and P02 across eight channels are shown for stimulation of the radial nerve (RN), median nerve (MN), and ulnar nerve (UN). For each nerve, waveforms from multiple sessions are overlaid to demonstrate the temporal consistency of the evoked potentials. Insets provide magnified views of the main response components. Time is shown in milliseconds and amplitude in microvolts.

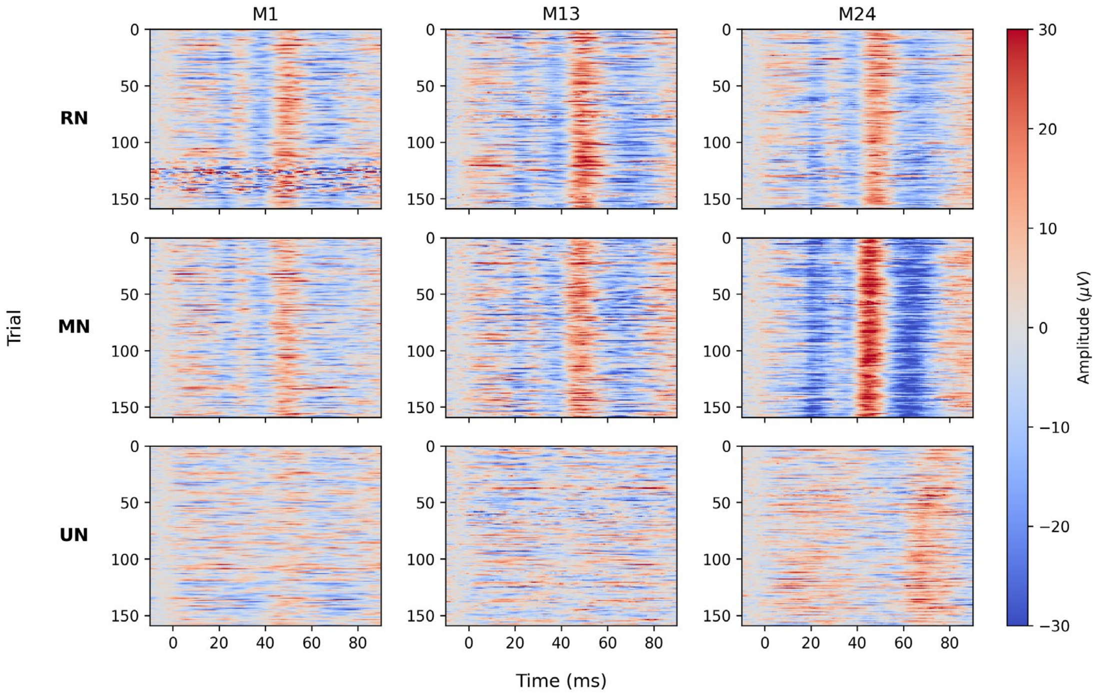

Figure S13 Longitudinal trial-by-trial maps of somatosensory evoked potentials for patient 01 (P01).

Single-trial response heat maps are shown for radial nerve (RN), median nerve (MN), and ulnar nerve (UN) stimulation at Month 1, Month 13, and Month 24. Each row corresponds to one trial, and color denotes amplitude in microvolts. Robust evoked responses were observed for radial and median nerve stimulation across all sessions, while responses to ulnar nerve stimulation were less prominent.

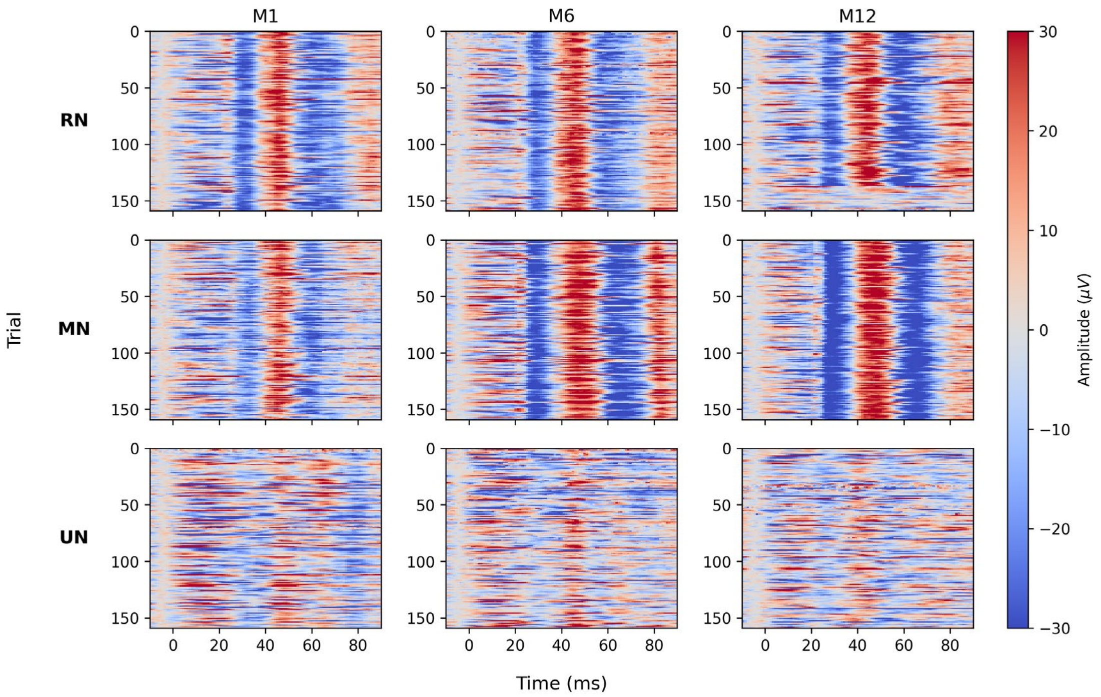

Figure S14 Longitudinal trial-by-trial somatosensory evoked responses in P02.

Heat maps show single-trial evoked responses recorded during stimulation of the radial nerve (RN), median nerve (MN), and ulnar nerve (UN) at M1, M6, and M12. Each row represents one trial, and color indicates signal amplitude (μV). Clear and stable evoked patterns were observed for radial and median nerve stimulation across sessions, whereas ulnar nerve responses were comparatively weaker.

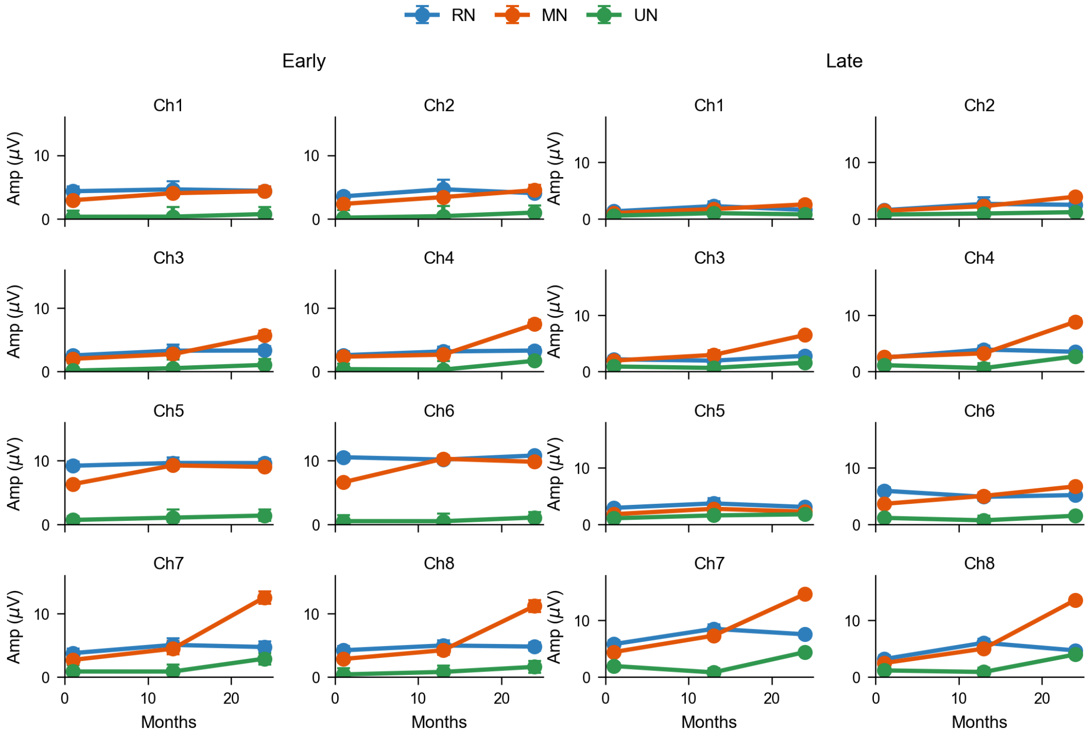

Figure S15 Changes in average SEP component amplitude over time of P01.

Average SEP amplitudes across months are shown for the **early component** (**15–35 ms**) and the **late component** (**40–80 ms**) for each recording channel. Within each subplot, responses to **radial nerve (RN)**, **median nerve (MN)**, and **ulnar nerve (UN)** stimulation are plotted separately. The x-axis indicates month, and the y-axis indicates amplitude (μV).

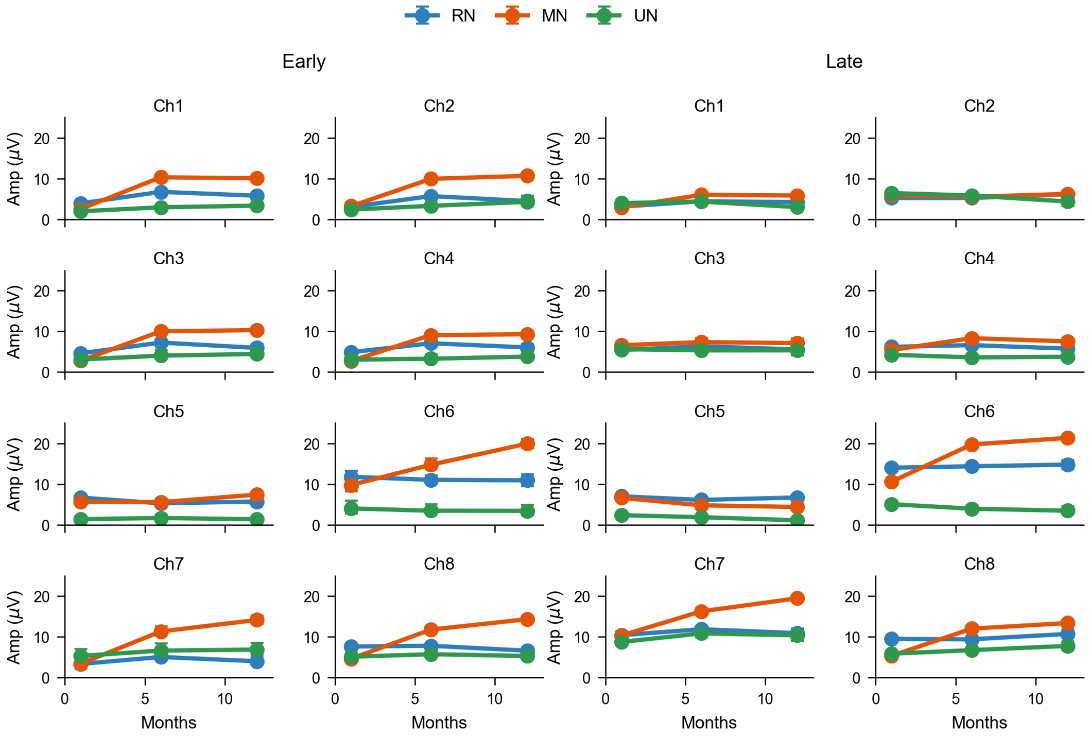

Figure S16 Changes in average SEP component amplitude over time of P02.

Average SEP amplitudes across months are shown for the early component (15–35 ms) and the late component (40–80 ms) for each recording channel. Within each subplot, responses to radial nerve (RN), median nerve (MN), and ulnar nerve (UN) stimulation are plotted separately. The x-axis indicates month, and the y-axis indicates amplitude (μV).

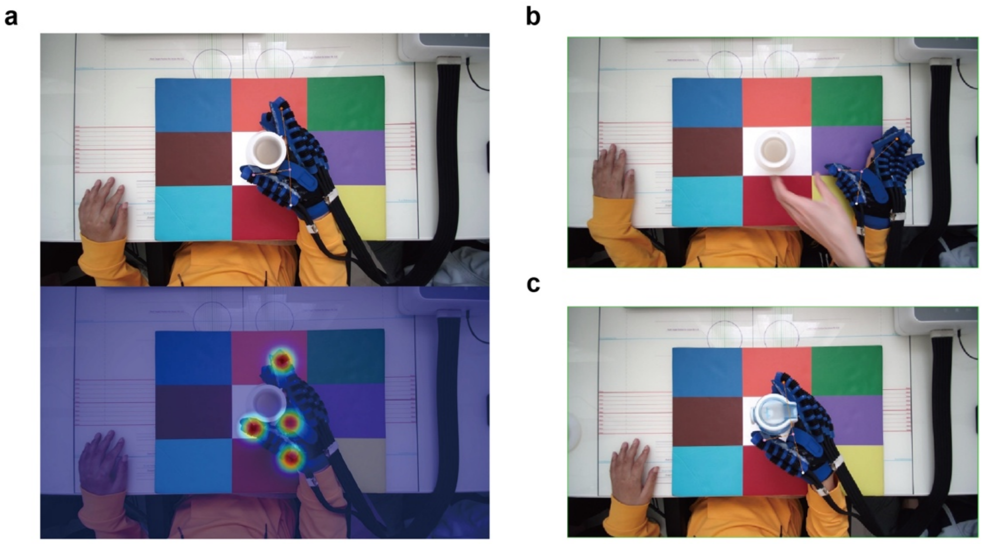

Figure S17 Pneumatic Hand Keypoint Detection Model.

a, Example of a detection heatmap. b-c, Examples of detection results.

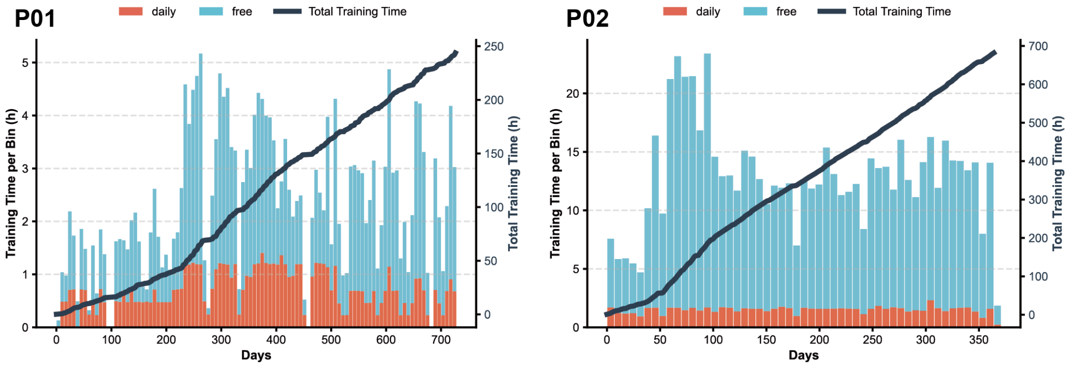

Figure S18 Training duration in the two participants for a combined 36 months BCI training.

Daily training time (blue bars, left y-axis) and cumulative total training time (dark line, right y-axis) are shown as a function of days for P01 (left) and P02 (right). The cumulative curves illustrate the progressive increase in total training exposure in both participants across the study. Each bin contains 7 days. Cue-locked daily evaluation sessions accounted for 30.1% and 11.6% of total training time in P01 and P02, respectively.

### Supplemental Tables

Table S1 ISNCSCI Score of P01

| Time | Pre-Operation (Day -4) | Month 3 (Day 78) | Month 6 (Day 162) | Month 7 (Day 198) | Month 8 (Day 234) | Month 9 (Day 279) | Month 12 (Day 367) | Month 28 (Day 841) |
|---|---|---|---|---|---|---|---|---|
| American Spinal Injury Association Impairment Scale (AIS) | A | A | A | A | A | A | A | A |
| Neurological level of injury | C4 | C4 | C4 | C4 | C4 | C4 | C4 | C4 |
| Motor Score | | | | | | | | |
| C5, (Left \| Right) | 4 \| 4 | 4 \| 4 | 4 \| 5 | 4 \| 5 | 4 \| 5 | **5** \| 5 | **5** \| 5 | **5** \| 5 |
| C6, (Left \| Right) | 3 \| 3 | 3 \| 3 | 3 \| **4** | 3 \| **3** | 3 \| 3 | 3 \| **4** | 3 \| **4** | 4 \| **4** |
| C7, (Left \| Right) | 2 \| 2 | 2 \| 2 | 2 \| **2** | 2 \| **2** | 2 \| **3** | 2 \| 3 | 2 \| 2 | 2 \| 2 |
| C8, (Left \| Right) | 0 \| 0 | 0 \| 0 | 0 \| 0 | 0 \| 0 | **1** \| 0 | **0** \| **1** | **1** \| **1** | **0** \| **0** |
| T1, (Left \| Right) | 0 \| 0 | 0 \| 0 | 0 \| 0 | 0 \| 0 | 0 \| 0 | 0 \| 0 | 0 \| 0 | 0 \| 0 |
| L2, (Left \| Right) | 0 \| 0 | 0 \| 0 | 0 \| 0 | 0 \| 0 | 0 \| 0 | 0 \| 0 | 0 \| 0 | 3 \| 4 |
| L3, (Left \| Right) | 0 \| 0 | 0 \| 0 | 0 \| 0 | 0 \| 0 | 0 \| 0 | 0 \| 0 | 0 \| 0 | 2 \| 3 |
| Total, (Left \| Right) | 9 \| 9 | 9 \| 9 | 9 \| 11 | 9 \| 10 | 10 \| 11 | 10 \| 13 | 11 \| 12 | 16 \| 18 |
| Light Touch Sensory Score | | | | | | | | |
| Total, (Left \| Right) | 7 \| 8 | 9 \| 7 | 8 \| 9 | 11 \| 9 | 8 \| 6 | 7 \| 7 | 7 \| 9 | 9 \| 10 |
| Pin Prick Sensory Score | | | | | | | | |
| Total, (Left \| Right) | 6 \| 7 | 10 \| 8 | 10 \| 8 | 11 \| 8 | 10 \| 10 | 11 \| 11 | 9 \| 12 | 11 \| 10 |
| Deep Anal Pressure | No | No | No | No | No | No | No | No |
| Voluntary Anal Contraction | No | No | No | No | No | No | No | No |

Table S2 ISNCSCI Score of P02

| Time | Pre-Operation (Day -1) | Month 2 (Day 67) | Month 6 (Day 198) |
|---|---|---|---|
| American Spinal Injury Association Impairment Scale (AIS) | B | C | C |
| Neurological level of injury | C7 | C7 | C7 |
| Upper Extremity Motor Score | | | |
| C5, (Left \| Right) | 5 \| 5 | 5 \| 5 | 5 \| 5 |
| C6, (Left \| Right) | 5 \| 5 | 5 \| 5 | 5 \| 5 |
| C7, (Left \| Right) | 5 \| 5 | 4 \| 4 | 5 \| 3 |
| C8, (Left \| Right) | 0 \| 0 | 1 \| 1 | 2 \| 2 |
| T1, (Left \| Right) | 0 \| 0 | 3 \| 1 | 3 \| 0 |
| Total, (Left \| Right) | 15 \| 15 | 18 \| 16 | 20 \| 15 |
| Light Touch Sensory Score | | | |
| Total, (Left \| Right) | 39 \| 40 | 40 \| 40 | 40 \| 49 |
| Pin Prick Sensory Score | | | |
| Total, (Left \| Right) | 38 \| 39 | 40 \| 40 | 40 \| 36 |
| Deep Anal Pressure | Yes | Yes | Yes |
| Voluntary Anal Contraction | No | Yes | Yes |

Table S3 p-values for SEP component changes in P01 and P02

| | | RN | | MN | | UN | |
|---|---|---|---|---|---|---|---|
| | | Early | Late | Early | Late | Early | Late |
| P01 | Ch1 | 0.4508 | 0.2751 | <0.0001 *** | <0.0001 *** | 0.168 | 0.2284 |
| | Ch2 | 0.1858 | 0.004 ** | <0.0001 *** | <0.0001 *** | 0.1 | 0.1166 |
| | Ch3 | 0.0904 | 0.0717 | <0.0001 *** | <0.0001 *** | 0.0496 * | 0.0166 * |
| | Ch4 | 0.0904 | 0.004 ** | <0.0001 *** | <0.0001 *** | 0.012 * | <0.0001 *** |
| | Ch5 | 0.2191 | 0.3685 | <0.0001 *** | 0.0303 ** | 0.1147 | 0.0512 |
| | Ch6 | 0.2744 | 0.9831 | <0.0001 *** | <0.0001*** | 0.1369 | 0.11656 |
| | Ch7 | 0.0904 | <0.0001 *** | <0.0001 *** | <0.0001*** | 0.004 ** | <0.0001 *** |
| | Ch8 | 0.1858 | <0.0001 *** | <0.0001 *** | <0.0001*** | 0.0328 * | <0.0001 *** |
| P02 | Ch1 | <0.0001*** | 0.0048 ** | <0.0001 *** | <0.0001 *** | 0.0192 * | 1.0 |
| | Ch2 | <0.0001*** | 0.0688 | <0.0001 *** | 0.2259 | 0.0192 * | 1.0 |
| | Ch3 | 0.0056 ** | 0.5485 | <0.0001 *** | 0.2259 | 0.0685 | 1.0 |
| | Ch4 | 0.01 * | 0.774 | <0.0001 *** | <0.0001 *** | 0.1324 | 1.0 |
| | Ch5 | 0.9899 | 0.774 | 0.003 ** | 1.0 | 0.5942 | 1.0 |
| | Ch6 | 0.9899 | 0.2752 | <0.0001 *** | <0.0001 *** | 0.7128 | 1.0 |
| | Ch7 | 0.329 | 0.4109 | <0.0001 *** | <0.0001 *** | 0.1715 | 0.0168 * |
| | Ch8 | 0.9899 | 0.0688 | <0.0001 *** | <0.0001 *** | 0.5642 | 0.0024 ** |

P-values for the comparison of early and late SEP components between the first and last months in P01 (M1 vs. M24) and P02 (M1 vs. M12) for each stimulated nerve. All p-values were FDR-corrected.

Table S4 BCI-assisted object transfer success rate for P01

| Session | Average time（s） | Success / Total |
|---|---|---|
| 1 | 5.45 | 16/16 |
| 2 | 4.86 | 17/17 |
| 3 | 5.33 | 16/16 |

Table S5 Unaided object transfer success rate for P01

| Session | Average time（s） | Success / Total |
|---|---|---|
| 1 | 5.09 | 5/16 |
| 2 | 4.22 | 8/16 |
| 3 | 7.91 | 4/16 |

Table S6 BCI-assisted object transfer success rate for P02

| Session | Average time（s） | Success / Total |
|---|---|---|
| 1 | 8.12 | 8/8 |
| 2 | 8.50 | 8/8 |
| 3 | 8.75 | 8/8 |
| 4 | 9.00 | 8/8 |
| 5 | 10.50 | 7/8 |
| 6 | 9.12 | 7/8 |

Table S7 ARAT Scores of P01

| | Baseline (Day 15) | | Month 3 (Day 77) | | Month 5 (Day 147) | | Month 7 (Day 204) | | Month 9 (Day 280) | | Month 12 (Day 367) | | Month 28 (Day 833) | |
|---|---|---|---|---|---|---|---|---|---|---|---|---|---|---|
| | **L** | **R** | **L** | **R** | **L** | **R** | **L** | **R** | **L** | **R** | **L** | **R** | **L** | **R** |
| **Grasp (18)** | 6 | 4 | 5 | 6 | 6 | 10 | 6 | 9 | 12 | 13 | 12 | 14 | 13 | 15 |
| **Grip (12)** | 6 | 6 | 6 | 6 | 6 | 6 | 6 | 6 | 7 | 10 | 7 | 10 | 7 | 10 |
| **Pinch (18)** | 2 | 2 | 2 | 2 | 4 | 4 | 3 | 4 | 5 | 6 | 5 | 8 | 5 | 9 |
| **Gross (9)** | 7 | 9 | 8 | 9 | 8 | 8 | 8 | 8 | 8 | 8 | 8 | 9 | 9 | 9 |
| **Total (57)** | 21 | 21 | 21 | 23 | 24 | 28 | 23 | 27 | 32 | 37 | 32 | 41 | 34 | 43 |

Table S8 ARAT Scores of P02

| | Baseline (Day -1) | | Month 2 (Day 67) | | Month 3 (Day 107) | | Month 4 (Day 141) | | Month 5 (Day 174) | | Month 6 (Day 200) | | Month 8 (Day 256) | | Month 10 (Day 311) | | Month 12 (Day 373) | |
|---|---|---|---|---|---|---|---|---|---|---|---|---|---|---|---|---|---|---|
| | **L** | **R** | **L** | **R** | **L** | **R** | **L** | **R** | **L** | **R** | **L** | **R** | **L** | **R** | **L** | **R** | **L** | **R** |
| **Grasp (18)** | 8 | 7 | 9 | 9 | 13 | 10 | 15 | 13 | 16 | 15 | 16 | 16 | 16 | 15 | 17 | 17 | 18 | 15 |
| **Grip (12)** | 6 | 4 | 9 | 9 | 9 | 9 | 10 | 9 | 10 | 9 | 10 | 9 | 10 | 9 | 11 | 9 | 10 | 7 |
| **Pinch (18)** | 4 | 2 | 4 | 0 | 5 | 0 | 5 | 0 | 5 | 1 | 6 | 2 | 7 | 2 | 7 | 2 | 8 | 3 |
| **Gross (9)** | 9 | 9 | 9 | 9 | 9 | 9 | 9 | 9 | 9 | 9 | 9 | 9 | 9 | 9 | 9 | 9 | 9 | 9 |
| **Total (57)** | 27 | 22 | 31 | 27 | 36 | 28 | 39 | 31 | 40 | 34 | 41 | 36 | 42 | 35 | 44 | 37 | 45 | 34 |

Table S9 Adverse Events reported during the 1-year follow-up period

| Screening No. | AE Term | Start Date | End Date | Pattern | Severity | SAE? | Relationship to Investigational Device | Outcome | Sequelae | Corrective Treatment | Measures Taken |
|---|---|---|---|---|---|---|---|---|---|---|---|
| P01 | Wound pain | 2023.10.25 | 2023.10.29 | Persistent | Mild | No | Possibly unrelated | Resolved | None | Yes | Flurbiprofen axetil injection 50 mg, once daily. |
| P01 | Cough | 2023.10.25 | 2023.11.02 | Persistent | Mild | No | Definitely unrelated | Resolved | None | Yes | Compound Fresh Bamboo Juice oral liquid, orally, 3 times daily. |
| P01 | Insomnia | 2023.10.25 | 2024.1.10 | Persistent | Mild | No | Definitely unrelated | Resolved | None | Yes | Estazolam tablets 2 mg orally (once nightly) and zolpidem tartrate tablets 10 mg orally (once nightly). |
| P01 | Constipation | 2023.10.26 | 2023.10.28 | Paroxysmal | Mild | No | Definitely unrelated | Resolved | None | Yes | Glycerin enema 40 mL with manual rectal assistance for defecation. |

### Reference

1. Fischl, B. FreeSurfer. *NeuroImage* **62**, 774–781 (2012).

2. Wang, J. *et al.* Deep High-Resolution Representation Learning for Visual Recognition. *IEEE Trans. Pattern Anal. Mach. Intell.* **43**, 3349–3364 (2021).

3. Zhang, F., Zhu, X., Dai, H., Ye, M. & Zhu, C. Distribution-Aware Coordinate Representation for Human Pose Estimation. *Proc. IEEE Comput. Soc. Conf. Comput. Vis. Pattern Recognit.* 7091–7100 (2020) doi:10.1109/CVPR42600.2020.00712.

4. Congedo, M., Barachant, A. & Bhatia, R. Riemannian geometry for EEG-based brain-computer interfaces; a primer and a review. *Brain-Comput. Interfaces* **4**, 155–174 (2017).

5. Yamamoto, M. S., Lotte, F., Yger, F. & Chevallier, S. Class-distinctiveness-based frequency band selection on the Riemannian manifold for oscillatory activity-based BCIs: preliminary results. in *2022 44th Annual International Conference of the IEEE Engineering in Medicine & Biology Society (EMBC)* vols 2022-July 3690–3693 (IEEE, 2022).

6. Barachant, A., Bonnet, S., Congedo, M. & Jutten, C. Classification of covariance matrices using a Riemannian-based kernel for BCI applications. *Neurocomputing* **112**, 172–178 (2013).

7. Benabid, A. L. *et al.* An exoskeleton controlled by an epidural wireless brain–machine

interface in a tetraplegic patient: a proof-of-concept demonstration. *Lancet Neurol.* **18**, 1112–1122 (2019).

8. Moly, A. *et al.* An adaptive closed-loop ECoG decoder for long-term and stable bimanual control of an exoskeleton by a tetraplegic. *J. Neural Eng.* **19**, 026021 (2022).

9. Koles, Z. J. The quantitative extraction and topographic mapping of the abnormal components in the clinical EEG. *Electroencephalogr. Clin. Neurophysiol.* **79**, 440–447 (1991).

10. Lemm, S., Blankertz, B., Curio, G. & Müller, K. R. Spatio-spectral filters for improving the classification of single trial EEG. *IEEE Trans. Biomed. Eng.* **52**, 1541–1548 (2005).

11. Miller, K. J. A library of human electrocorticographic data and analyses. *Nat. Hum. Behav.* **3**, 1225–1235 (2019).

12. Lotze, M. & Halsband, U. Motor imagery. *J. Physiol. Paris* **99**, 386–395 (2006).

13. Schalk, G., McFarland, D. J., Hinterberger, T., Birbaumer, N. & Wolpaw, J. R. BCI2000: A General-Purpose Brain-Computer Interface (BCI) System. *IEEE Trans. Biomed. Eng.* **51**, 1034–1043 (2004).

14. Kobak, D. *et al.* Demixed principal component analysis of neural population data. *eLife* **5**, 1–36 (2016).

15. Williams, A. H. *et al.* Unsupervised Discovery of Demixed, Low-Dimensional Neural Dynamics across Multiple Timescales through Tensor Component Analysis. *Neuron* **98**,

1099-1115.e8 (2018).

16. Cruccu, G. *et al.* Recommendations for the clinical use of somatosensory-evoked potentials. *Clin. Neurophysiol.* **119**, 1705–1719 (2008).

17. Allison, T., McCarthy, G., Wood, C. C. & Jones, S. J. Potentials evoked in human and monkey cerebral cortex by stimulation of the median nerve. A review of scalp and intracranial recordings. *Brain* **114**, 2465–2503 (1991).

18. Allison, T., McCarthy, G. & Wood, C. C. The relationship between human long-latency somatosensory evoked potentials recorded from the cortical surface and from the scalp. *Electroencephalogr. Clin. Neurophysiol. Evoked Potentials* **84**, 301–314 (1992).

19. Jorge, A. *et al.* Area under the Curve of Somatosensory Evoked Potentials Detects Spinal Cord Injury. *J. Clin. Neurophysiol.* **36**, 155–160 (2019).

20. Rupp, R. *et al.* International standards for neurological classification of spinal cord injury. *Top. Spinal Cord Inj. Rehabil.* **27**, 1–22 (2021).

21. Yozbatiran, N., Der-Yeghiaian, L. & Cramer, S. C. A standardized approach to performing the action research arm test. *Neurorehabil. Neural Repair* **22**, 78–90 (2008).

22. Balaguer, J.-M. *et al.* Neural mechanisms underlying the recovery of voluntary control of motoneurons after paralysis with spinal cord stimulation. *Neuron* **113**, 3846-3862.e6 (2025).

23. Dan, Y. & Poo, M. Spike Timing-Dependent Plasticity of Neural Circuits. *Neuron* **44**,

23–30 (2004).

24. Feldman, D. E. The Spike-Timing Dependence of Plasticity. *Neuron* **75**, 556–571 (2012).

25. Capogrosso, M. *et al.* A Computational Model for Epidural Electrical Stimulation of Spinal Sensorimotor Circuits. *J. Neurosci.* **33**, 19326–19340 (2013).

26. Fuglevand, A. J., Winter, D. A. & Patla, A. E. Models of recruitment and rate coding organization in motor-unit pools. *J. Neurophysiol.* **70**, 2470–2488 (1993).
